# Evaluating Individual Level Performance of Polygenic Risk Scores Using Early Onset High Genetic Risk Coronary Artery Disease as a Benchmark

**DOI:** 10.64898/2026.04.16.26350801

**Authors:** Shengxin Liang, Min Seo Kim, Yang Sui, Youxin Tan, Linke Li, So Mi Jemma Cho, Satoshi Koyama, Yixuan Liu, Kaavya Paruchuri, Andrew Chan, Michael C. Honigberg, Pradeep Natarajan, Nilanjan Chatterjee, Akl C. Fahed, Zhi Yu

## Abstract

Polygenic risk scores (PRSs) are typically validated using population-level metrics, masking variability in individual-level risk prediction and hindering clinical translation. To address this, we introduced a novel framework using a “benchmark” cohort (N=1184) of “unexpected coronary artery disease (CAD)”: early-onset patients (<55 years) with a clinical profile—low 10-year risk, no diabetes or severe hypercholesterolemia—that excludes therapy indications. The occurrence of early CAD in these clinically low-risk individuals establishes a “ground truth” for high genetic risk. We evaluated 58 published CAD PRSs and demonstrated a disconnection between population-level performance and individual-level accuracy (proportion of benchmark patients captured). The proportion captured by 58 PRSs varied from 10.8% to 33.1%, and the top-performing score was 2-fold more effective at identifying the benchmark group than established non-genetic biomarkers, such as lipoprotein(a). Furthermore, benchmark patients never captured by any score exhibited significantly healthier lipid profiles. Our framework provides an essential method for validating clinical readiness of PRSs.

## Introduction

Polygenic risk scores (PRSs) aggregate the effects of risk variants to summarize the genetic components of a trait and have been proposed as a tool for risk prediction^1^. While the field has excelled at developing PRSs optimized for aggregate metrics such as area under the curve (AUC) or variance explained (r^2^)^2,3^, the clinical translation^4^ of PRSs is hampered by a disconnection between performance at the population level and utility for individual patient assessment^4–6^. This gap was starkly illustrated in recent studies, which found that dozens of coronary artery disease (CAD) PRSs with practically identical population-level performances yielded highly variable and poorly correlated risk estimates for the same individual^5,6^. This incongruence undermines clinical confidence and creates an urgent need for new evaluation methods that directly test the performance of a PRS where it matters most: at the level of individual patients with the highest genetic risks.

To address this challenge, we introduced a new framework to benchmark the individual-level accuracy of PRS using “unexpected CAD” as a critical test case. We defined this benchmark group as individuals who developed early-onset CAD despite meeting the clinical criteria for low risk—specifically, low Pooled Cohort Equation (PCE)-based 10-year risk, no diabetes mellitus, and no severe hypercholesterolemia. As this specific profile excludes the indication for lipid-lowering therapy under current guidelines, the occurrence of early-onset disease in this group serves as a potent indicator of high underlying genetic predisposition. In addition, we curated a more stringent set of unexpected CAD cases for sensitivity analysis, consisting of individuals devoid of conventional drivers (i.e., smoking, diabetes mellitus, hypertension and hyperlipidemia)^7^, making it even more likely attributable to a high burden of genetic risk. We posited that any clinically useful PRSs must be capable of identifying a significant proportion of these benchmark individuals. This benchmark thus provided a stringent and interpretable test of a PRS’s fundamental purpose: identifying individuals truly at high genetic risk.

In this study, we developed and applied our benchmark framework to systematically evaluate all 58 published CAD PRSs in UK Biobank (UKBB). We first assessed the capacity of each score to identify the benchmark population (early onset, unexpected CAD). We then contrasted this direct measure of individual-level accuracy against conventional population-level performance metrics to systematically investigate discordance between these two evaluation paradigms. Finally, we explored how PRSs performed relative to other clinical biomarkers and how their performances varied across ancestral backgrounds. Our study aims to provide a robust and transparent methodology for evaluating the individual-level accuracy of any given PRS, thereby contributing to the broader effort of translating PRS from population-level association to meaningful individual-level prediction.

## Results

### Population Characteristics

For our primary analysis, we identified 41,397 individuals in UKBB with prevalent or incident CAD and complete risk factor information. CAD cases were defined using a combination of ICD9, ICD10, and OPCS-4 (Office of Population Censuses and Surveys Classification of Interventions and Procedures version 4) codes, as detailed in **Supplementary Table 1**. We stratified this cohort into two groups based on their clinical risk profiles. We defined “unexpected CAD” by the absence of standard indications for lipid-lowering therapy: specifically, a low PCE-based 10-year risk (<5%)^8^, absence of diabetes (hemoglobin A1c [HbA1c] < 6.5%, no diabetes mellitus medication), and no severe hypercholesterolemia (low-density lipoprotein cholesterol [LDL-C] < 4.9 mmol/L^9^, no lipid-lowering medication). By excluding individuals taking lipid-lowering or antidiabetic medications at the time of their baseline measurement, the biomarker values utilized in our analysis strictly reflect an unmedicated physiological state to prevent potential reverse causation or misclassification. Because the disease etiology in this group cannot be explained by non-genetic risk factors, their phenotype implies a dominant genetic component.

This stratification yielded 3,782 individuals with unexpected CAD and 37,615 individuals categorized as “expected” CAD (driven by conventional risk factors). Compared with the expected group, the unexpected CAD cohort was younger, had a smaller proportion of males and British White population, and a higher level of socioeconomic deprivation (reflected by Townsend Deprivation Index [TDI]). As anticipated, this group exhibited a significantly healthier clinical profile, characterized by a higher (more favorable) overall Life’s Essential 8 (LE8) composite score, higher scores across most individual LE8 components (except for physical activity and sleep health)^10^, and fewer familial hypercholesterolemia genetic variant carriers (**Table 1**).

**Table 1.**
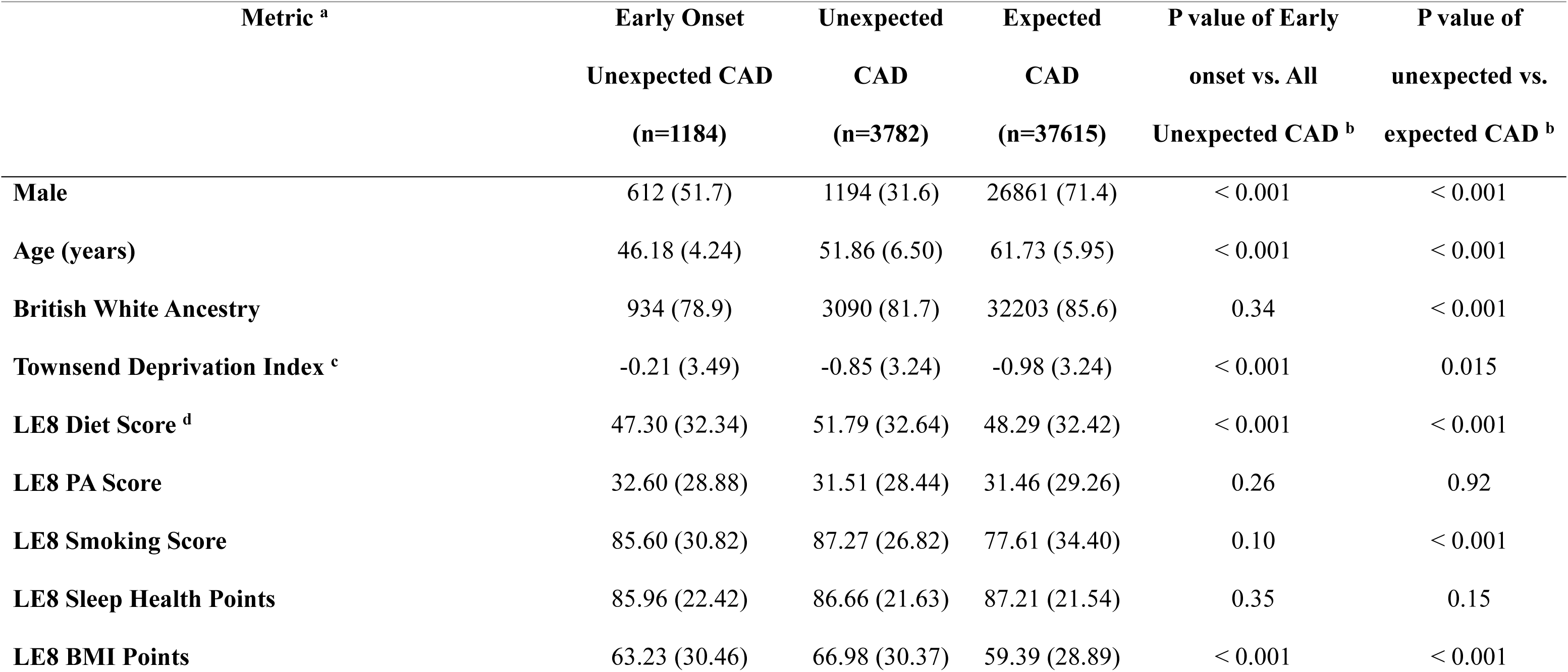
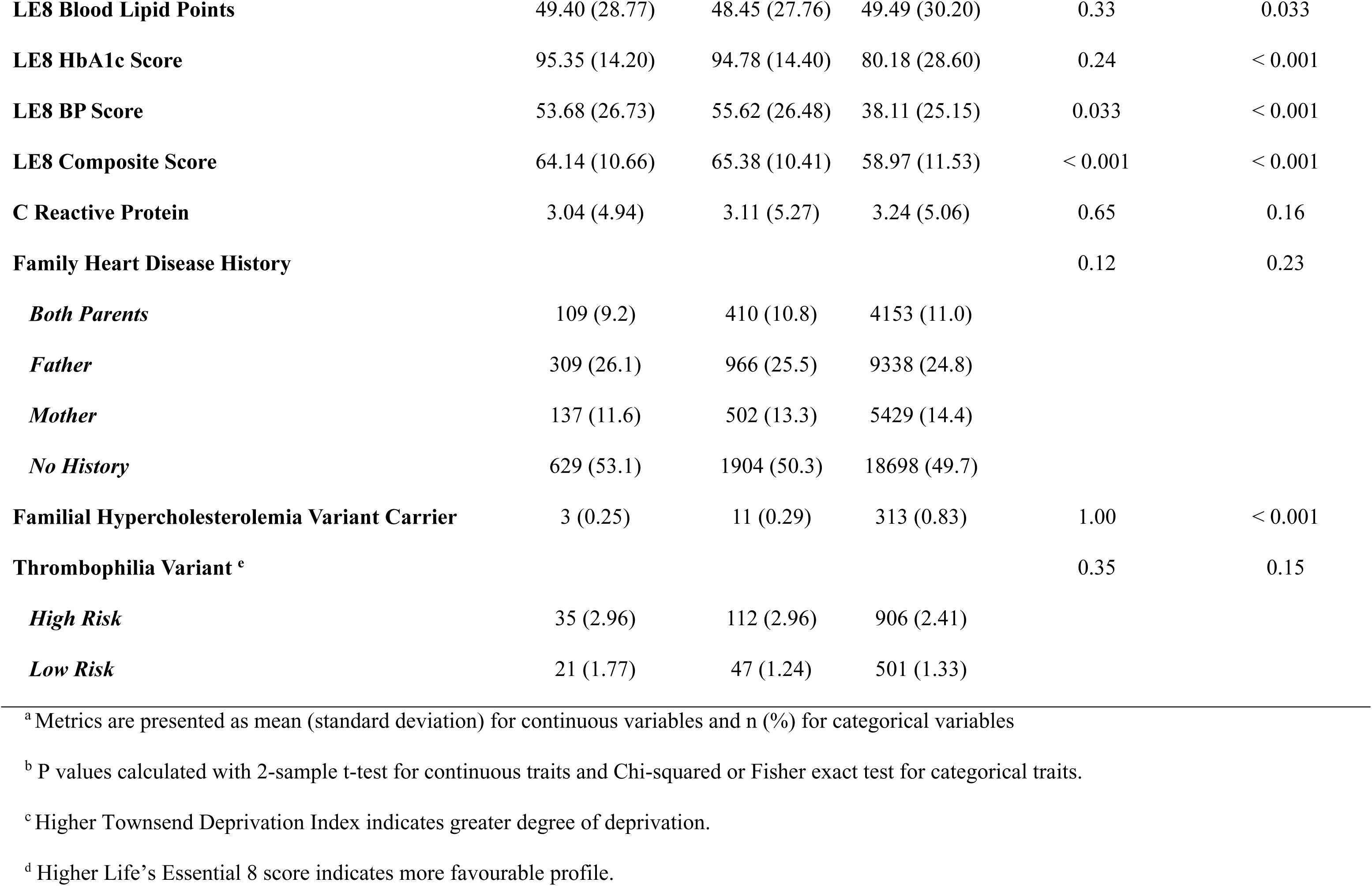
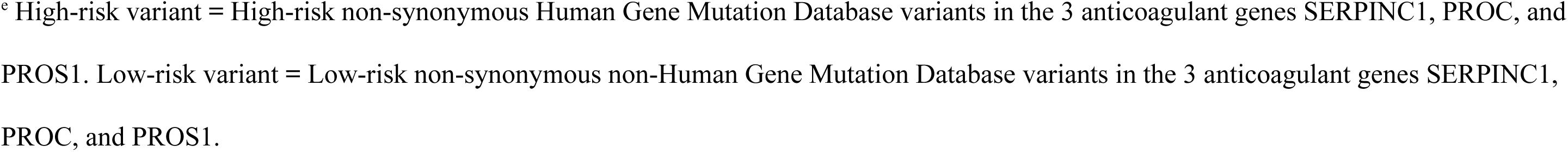
Baseline characteristics of UK Biobank participants with early onset unexpected coronary artery disease (CAD), unexpected CAD and expected CAD. Unexpected CAD was defined as CAD patients with low 10-year risk (Pooled Cohort Equation [PCE] risk < 0.05), absence of diabetes (hemoglobin A1c [HbA1c] < 6.5%, no diabetes mellitus medication) and no severe hypercholesterolemia (low-density lipoprotein cholesterol [LDL-C] < 4.9 mmol/L, no lipid-lowering medication). Expected CAD was defined as those having CAD and complete data on definitive criteria, but not qualified as unexpected CAD. Early onset age cutoff is 55 years old.

| **Metric <sup>a</sup>** | **Early Onset**<br>**Unexpected CAD**<br>**(n=1184)** | **Unexpected**<br>**CAD**<br>**(n=3782)** | **Expected**<br>**CAD**<br>**(n=37615)** | **P value of Early**<br>**onset vs. All**<br>**Unexpected CAD <sup>b</sup>** | **P value of**<br>**unexpected vs.**<br>**expected CAD <sup>b</sup>** |
| --- | --- | --- | --- | --- | --- |
| **Male** | 612 (51.7) | 1194 (31.6) | 26861 (71.4) | < 0.001 | < 0.001 |
| **Age (years)** | 46.18 (4.24) | 51.86 (6.50) | 61.73 (5.95) | < 0.001 | < 0.001 |
| **British White Ancestry** | 934 (78.9) | 3090 (81.7) | 32203 (85.6) | 0.34 | < 0.001 |
| **Townsend Deprivation Index <sup>c</sup>** | -0.21 (3.49) | -0.85 (3.24) | -0.98 (3.24) | < 0.001 | 0.015 |
| **LE8 Diet Score <sup>d</sup>** | 47.30 (32.34) | 51.79 (32.64) | 48.29 (32.42) | < 0.001 | < 0.001 |
| **LE8 PA Score** | 32.60 (28.88) | 31.51 (28.44) | 31.46 (29.26) | 0.26 | 0.92 |
| **LE8 Smoking Score** | 85.60 (30.82) | 87.27 (26.82) | 77.61 (34.40) | 0.10 | < 0.001 |
| **LE8 Sleep Health Points** | 85.96 (22.42) | 86.66 (21.63) | 87.21 (21.54) | 0.35 | 0.15 |
| **LE8 BMI Points** | 63.23 (30.46) | 66.98 (30.37) | 59.39 (28.89) | < 0.001 | < 0.001 |
| **LE8 Blood Lipid Points** | 49.40 (28.77) | 48.45 (27.76) | 49.49 (30.20) | 0.33 | 0.033 |
| **LE8 HbA1c Score** | 95.35 (14.20) | 94.78 (14.40) | 80.18 (28.60) | 0.24 | < 0.001 |
| **LE8 BP Score** | 53.68 (26.73) | 55.62 (26.48) | 38.11 (25.15) | 0.033 | < 0.001 |
| **LE8 Composite Score** | 64.14 (10.66) | 65.38 (10.41) | 58.97 (11.53) | < 0.001 | < 0.001 |
| **C Reactive Protein** | 3.04 (4.94) | 3.11 (5.27) | 3.24 (5.06) | 0.65 | 0.16 |
| **Family Heart Disease History** |  |  |  | 0.12 | 0.23 |
| *Both Parents* | 109 (9.2) | 410 (10.8) | 4153 (11.0) |  |  |
| *Father* | 309 (26.1) | 966 (25.5) | 9338 (24.8) |  |  |
| *Mother* | 137 (11.6) | 502 (13.3) | 5429 (14.4) |  |  |
| *No History* | 629 (53.1) | 1904 (50.3) | 18698 (49.7) |  |  |
| **Familial Hypercholesterolemia Variant Carrier** | 3 (0.25) | 11 (0.29) | 313 (0.83) | 1.00 | < 0.001 |
| **Thrombophilia Variant <sup>e</sup>** |  |  |  | 0.35 | 0.15 |
| *High Risk* | 35 (2.96) | 112 (2.96) | 906 (2.41) |  |  |
| *Low Risk* | 21 (1.77) | 47 (1.24) | 501 (1.33) |  |  |

<sup>a</sup> Metrics are presented as mean (standard deviation) for continuous variables and n (%) for categorical variables
<sup>b</sup> P values calculated with 2-sample t-test for continuous traits and Chi-squared or Fisher exact test for categorical traits.
<sup>c</sup> Higher Townsend Deprivation Index indicates greater degree of deprivation.
<sup>d</sup> Higher Life's Essential 8 score indicates more favourable profile.
<sup>e</sup> High-risk variant = High-risk non-synonymous Human Gene Mutation Database variants in the 3 anticoagulant genes SERPINC1, PROC, and PROS1. Low-risk variant = Low-risk non-synonymous non-Human Gene Mutation Database variants in the 3 anticoagulant genes SERPINC1, PROC, and PROS1.

To isolate a benchmark cohort with a disease profile most strictly attributable to genetic burden, we further restricted the unexpected CAD group to individuals with early-onset disease (age of onset <55 years). This final “early-onset, unexpected CAD” benchmark group (n =1,184) formed the core of our analysis. Compared to the broader unexpected CAD group, the benchmark population was younger, had a higher proportion of males, greater deprivation, and even more favorable LE8 risk scores (**Table 1**).

### Characteristics of CAD PRSs

We systematically evaluated all 58 CAD PRSs currently available in the PGS Catalog^11,12^, extracting detailed methodological and population data from both the catalog and source publications. To facilitate comparison, we categorized scores based on their development strategy: (1) Multi-ancestry scores, which utilized variant associations or development cohorts spanning more than one ancestry; (2) Multi-trait scores, which incorporated genetically correlated traits or CAD risk factors; and (3) Multi-method scores, which aggregated multiple distinct scores for the same trait. These categories were not mutually exclusive.

Of the 58 CAD PRSs available in the PGS Catalog, four used multi-trait methods (PGS002262: metaPRS^13^; PGS004513 & PGS004514: RFDiseasemetaPRS^14^; PGS004746: PRSmixPlus^15^) and four were multi-method (PGS000018: metaGRS^16^; PGS004743 & PGS004744 & PGS004745: PRSmix^15^). Thirty-five were categorized as multi-ancestry. Among the ancestry-specific scores for which a source population (used for variant association or score development) was reported, all were European-derived except for two South Asian scores^15^.

**Supplementary Table 2** summarizes population-level performance ranking based on odds ratio (OR) per standard deviation (SD), individual-level performance ranking based on proportion of benchmark population (early onset, unexpected CAD) captured in the top decile of the PRS distribution in UKBB, development details from PGS catalog^11,12^ (https://www.pgscatalog.org/), and source publication for each score.

### Sensitivity of PRSs in Identifying the Benchmark Cohort

To evaluate the individual-level risk prediction of PRSs, we calculated the sensitivity of each PRS—specifically, the proportion of benchmark patients (early-onset, unexpected CAD) falling within the top decile (top 10%) of the PRS distribution in UKBB. The decision to use the top decile as a high PRS cutoff was evaluated using a precision-recall curve (**Supplementary Figure 1**), where the threshold with considerable recall (> 0.3) and highest precision was chosen. **Figure 1** displays the individual-level performance across the entire UKBB population with 95% confidence interval (CI) from bootstrapping, as well as performance stratified by ancestry (British White vs. non-British White).

**Figure 1.**
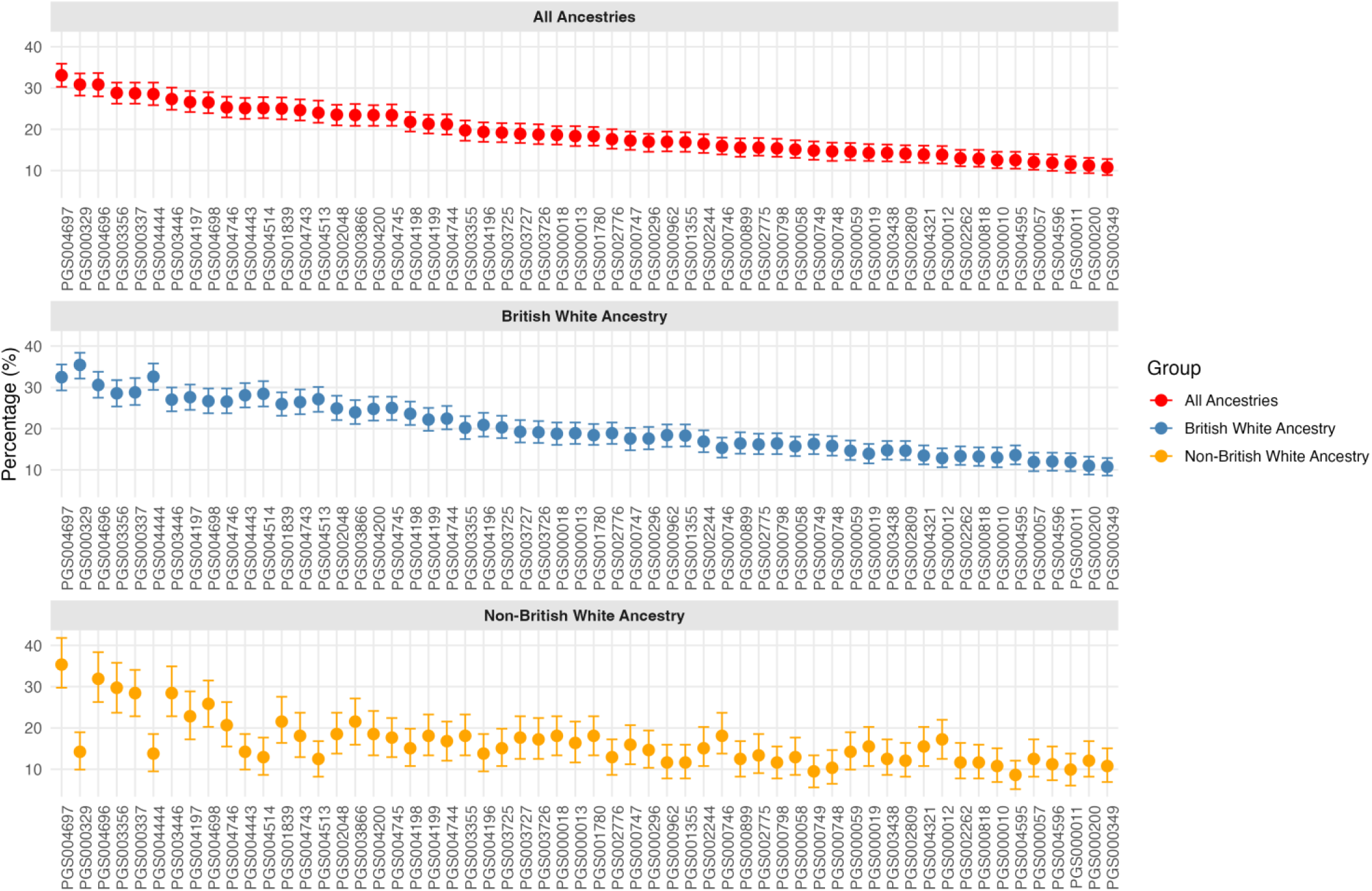
Individual-level risk prediction performance of 58 CAD PRS scores. Individual-level risk prediction of each PRS was measured by sensitivity, defined as the proportion of the benchmark population (n=1,184; individuals with early-onset, unexpected CAD) who fell within the top decile of each PRS distribution in the full UKBB population (n=451,580). The denominator is the total benchmark population (n=1,184), not the top PRS decile. For an uninformative PRS, approximately 10% of the benchmark group would be expected to fall in the top decile by chance alone; values exceeding 10% indicate enrichment of high genetic risk in the benchmark cohort. To move PRS performance test to absolute space, we evaluated the percentage of the benchmark population whose disease risk is unlikely to be explained by conventional factors and is therefore most plausibly attributable to genetic or inherited risk. We assumed that a PRS capturing a greater proportion of this benchmark population more effectively identifies individuals whose disease is genetically driven, thereby reflecting better individual-level predictive performance. The scores were ranked by decreasing percentage capture in all ancestries. Score percentage capture was also evaluated in British White and non-British White ancestry subsets. Error bars represent 95% confidence intervals obtained via bootstrapping.

We observed substantial variability in individual-level performances across the 58 CAD PRSs. The proportion of the benchmark (early-onset, unexpected CAD) group captured in the top PRS decile varied 3-fold, from 10.8% to 33.1% (**Figure 1**, ‘All Ancestries’). With a smaller sample size in the non-British White Ancestry subgroup (n=250), CIs are wider than those of the all ancestries or British White Ancestry analysis, but are still adequately powered to detect the large difference observed between the top vs. the lowest performing PRS pair. The highest performance was achieved by PGS004697^17^ (PRS-CSx^18^, multi-ancestry data for source of variant associations), followed by PGS004696^17^, PGS000329^19^, and PGS003356^20^, all of which were genome-wide scores using Bayesian approaches.

Performance consistency varied significantly by ancestry. As expected, given the demographic composition of the UKBB (predominantly British White participants), rankings in the British White cohort closely mirrored the overall “All Ancestries” ranking. The multi-ancestry scores (e.g., PGS004697^17^, PGS004696^17^, PGS003356^20^) showed, as expected, more consistent performances across both British White and non-British White populations; and ancestry-specific scores (e.g., PGS000329^19^, PGS004444^14^, PGS004197^21^), including those belonging to multi-trait or multi-method categories, exhibited a steep decline in their performance when evaluated in the non-British White population (**Figure 1**).

### Sensitivity of PRS versus Established Clinical Biomarkers

To contextualize the clinical utility of the PRS, we compared the performance of the top-scoring PRS (PGS004697^17^) against three established cardiovascular biomarkers: BMI, lipoprotein(a) [Lp(a)], and C-reactive protein (CRP). Acknowledging that practical clinical decisions rely on integrated models rather than binary thresholds of a single clinical biomarker, we also compared the top-scoring PRS with risk-estimating combinations, including PRS with Lp(a) and PRS with PCE. We used the top-scoring PRS and standardized Lp(a) in deriving the combinatory scores and performed McNemar’s test to evaluate differences in proportions of benchmark cohort (early onset, unexpected CAD) identified by different variable pairs (**Figure 2**, **Table 2**).

**Figure 2.**
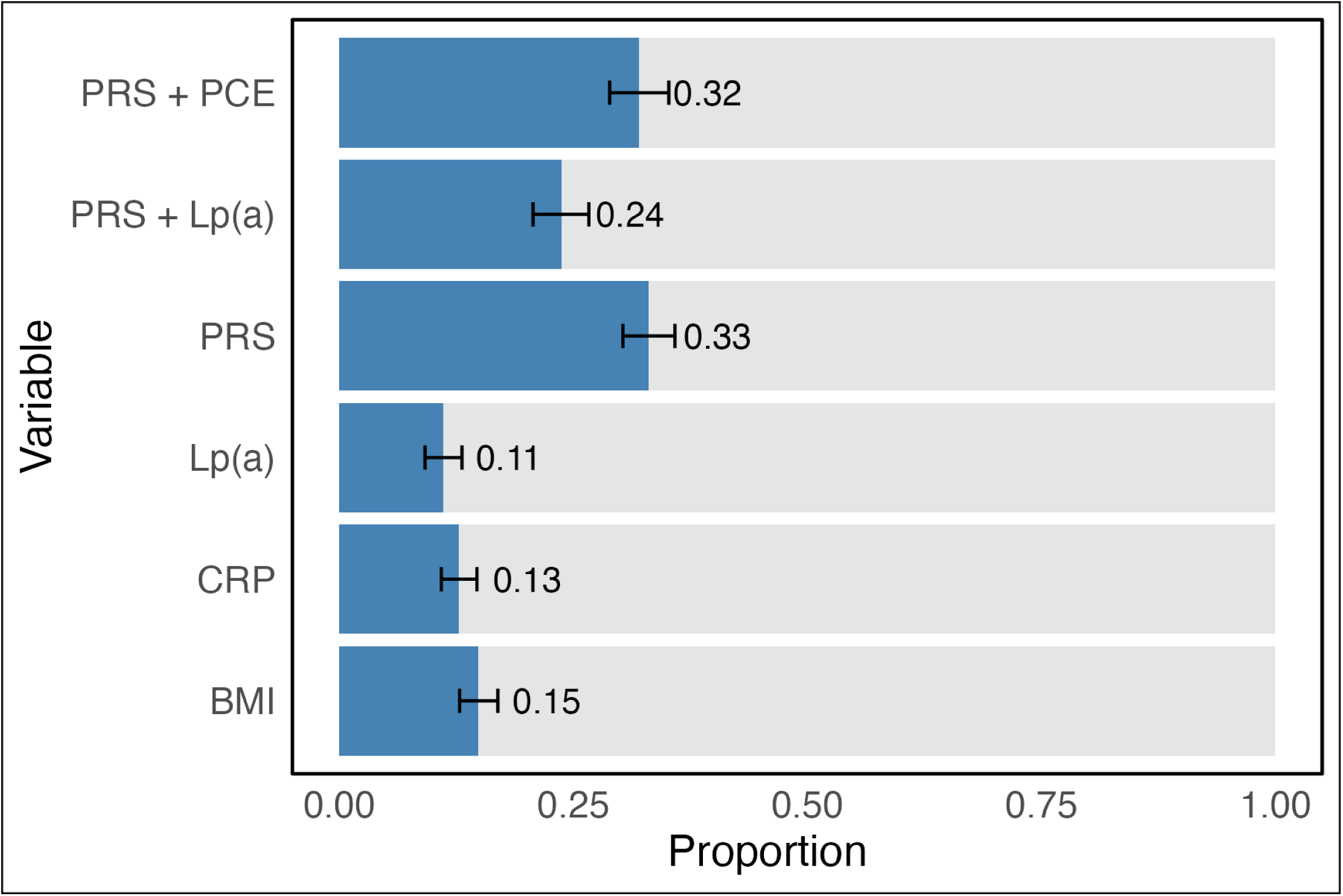
Proportion of early onset, unexpected CAD patients captured at the top decile of PRS versus other clinical biomarkers. The PRS used for comparison in this figure is the best-performing PRS among the 58 CAD PRS evaluated. Error bars represent 95% confidence intervals obtained via bootstrapping. Lp(a) was standardized before combining with PRS. BMI = body mass index; CRP = C-reactive protein; Lp(a) = lipoprotein (a); PCE = Pooled Cohort Equation; PRS = polygenic risk score.

**Table 2.** Proportion of early onset, unexpected CAD patients captured at the top decile of PRS versus other clinical biomarkers. Performances of a range of variables were compared with the top PRS (PGS004697). Median PRS was the individual-level median-performing PGS000013, while low PRS was the individual-level lowest-performing PGS000349. Lp(a) was standardized before combining with PRS. 95% confidence intervals were obtained via bootstrapping. BMI = body mass index; CRP = C-reactive protein; Lp(a) = lipoprotein (a); PCE = Pooled Cohort Equation; PRS = polygenic risk score. P-value was calculated with McNemar’s test.

| **Variable** | **Proportion [95% CI]** | **P-value <sup>a</sup>** |
| --- | --- | --- |
| **Top PRS** | 0.33 [0.30, 0.36] | Reference |
| **Median PRS** | 0.18 [0.16, 0.21] | < 0.01 |
| **Low PRS** | 0.11 [0.09, 0.13] | < 0.01 |
| **BMI** | 0.15 [0.13, 0.17] | < 0.01 |
| **Lp(a)** | 0.11 [0.09, 0.17] | < 0.01 |
| **CRP** | 0.13 [0.11, 0.15] | < 0.01 |
| **PRS + Lp(a)** | 0.24 [0.21, 0.27] | < 0.01 |
| **PRS + PCE** | 0.32 [0.29, 0.35] | 0.162 |

<sup>a</sup> P values calculated with McNemar's test.

The top PRS significantly outperformed conventional biomarkers, capturing 0.33 (95% CI: 0.30, 0.36) of the benchmark group in its top decile. This sensitivity was almost three-fold and statistically significantly higher than Lp(a) (0.11 [95% CI: 0.09, 0.13]), CRP (0.12 [95% CI: 0.11, 0.15]), and BMI (0.15 [95% CI: 0.13, 0.17]) (**Table 2**). The lowest- and median-performing PRS had comparable performances with the above clinical risk factors, evident from overlapping CIs (**Figure 2**). On the other hand, PRS as an addition to Lp(a) and PCE resulted in higher performance, at 0.24 (95% CI: 0.21, 0.27) and 0.32 (95% CI: 0.29, 0.35), respectively, highlighting the importance of identifying a top-performing PRS by our individual-level benchmark capture metric for clinical application.

### Population-Level Performance Across PRSs

To contrast individual-level accuracy with traditional metrics, we evaluated the population-level performance of all 58 scores using OR of CAD per SD increase in PRS. We used logistic regression here because the clinical factors used for defining this population were all measured at enrollment. The model was adjusted for age, sex, and the first four principal components of genetic ancestry (**Methods**). As in the individual-level analysis, we ranked scores based on the total UKBB population and performed stratified evaluations in British White and non-British White subgroups to assess cross-ancestry consistency (**Figure 3**).

**Figure 3.**
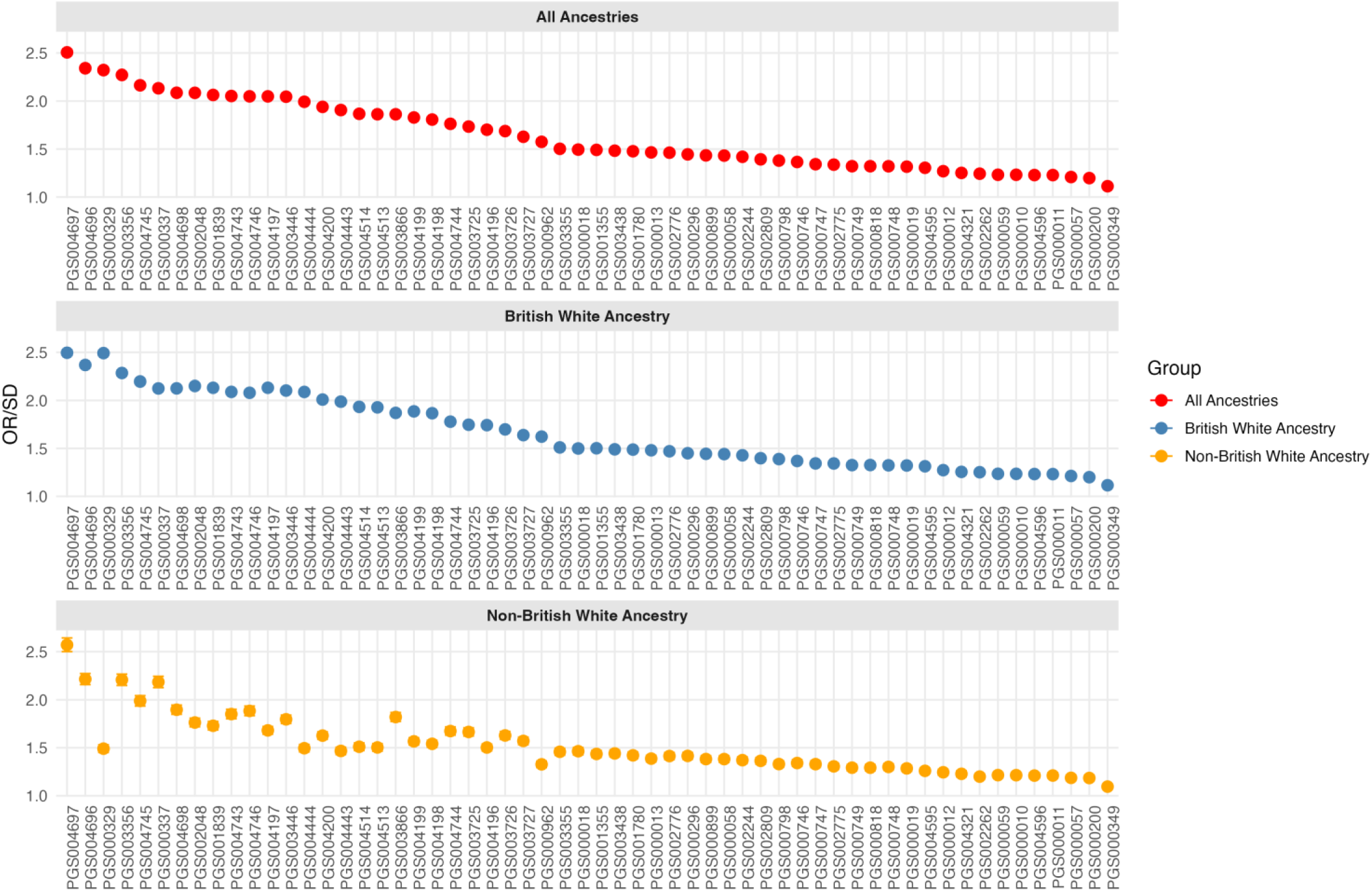
Population-level performance of 58 CAD PRS. Population-level performance of PRS scores was conventionally assessed with odds ratio (OR) per one standard deviation increase in PRS using a logistic regression model in UKBB population. The model was adjusted for age, sex and PC1-PC4. The scores were ranked by decreasing OR in all ancestries. Score OR was also evaluated in British White and non-British White ancestry separately. Error bars represent 95% confidence intervals obtained via bootstrapping.

Population-level performance varied widely, with ORs per SD ranging from 1.11 to 2.51. The top-performing scores mirrored the individual-level results: PGS004697^17^ (PRS-CSx^18^, multi-ancestry data for source of variant associations), followed by three other Bayesian methods-derived scores: PGS004696^17^, PGS000329^19^, and PGS003356^20^. As observed with the individual-level analysis, multi-ancestry scores demonstrated more concordant performances across ancestry groups, while ancestry-specific scores had marked decreases in performances in the non-British White population. However, the performance gap in the non-British White population—both in terms of absolute drop and ranking inconsistency—was significantly less pronounced in the population-level analysis (**Figure 3**) than in the individual-level benchmark (**Figure 1**).

### Concordance and Discordance in PRS Rankings

The rankings of population- and individual-level performance are correlated but exhibit notable differences (Spearman’s rho: 0.96; median ranking change [interquartile range]: -0.5 [4]). Among the scores ranked top 10 by individual-level performance, PGS004697^17^, PGS004696^17^, PGS000329^19^, PGS003356^20^ were genome-wide scores developed using Bayesian approaches (LDpred^22^, PRS-CSx^18^). These scores also ranked within top 10 by population-level performance, suggesting that Bayesian methods^23^ yielded relatively robust results across both evaluation metrics. In contrast, the remaining top-ranking scores at the individual level (PGS000337^24^, PGS003446^25^, PGS004197^21^, and PGS004698^17^) were constructed using methods that involve variant selection via pruning and thresholding (P+T)^26^ (e.g. PRSice^27^) or penalized regression (e.g., LASSO^21^). With the exception of PGS004698^17^, these scores had lower population-level rankings, moving an average of 5 places down (**Figure 4, Supplementary Table 2**). Multi-trait and multi-method scores were not among the top performed scores based on individual-level metrics. This is likely because incorporating data from other disease traits or aggregating multiple methods may dilute the specific signal required to identify high-risk individuals for a specific disease phenotype and population (**Figure 4, Supplementary Table 2**). Besides ranking changes, **Supplementary Figure 2** visualizes the substantial differences between individual-level performances of population-wise comparable scores comparing the number of benchmark cohort individuals identified across PRS deciles of the top and the worst individual-level performing PRSs. While individuals of the benchmark cohort were distributed nearly uniformly across the range of the worst-performing PRS, the top-performing PRS showed enrichment of the benchmark cohort in the top decile.

**Figure 4.**
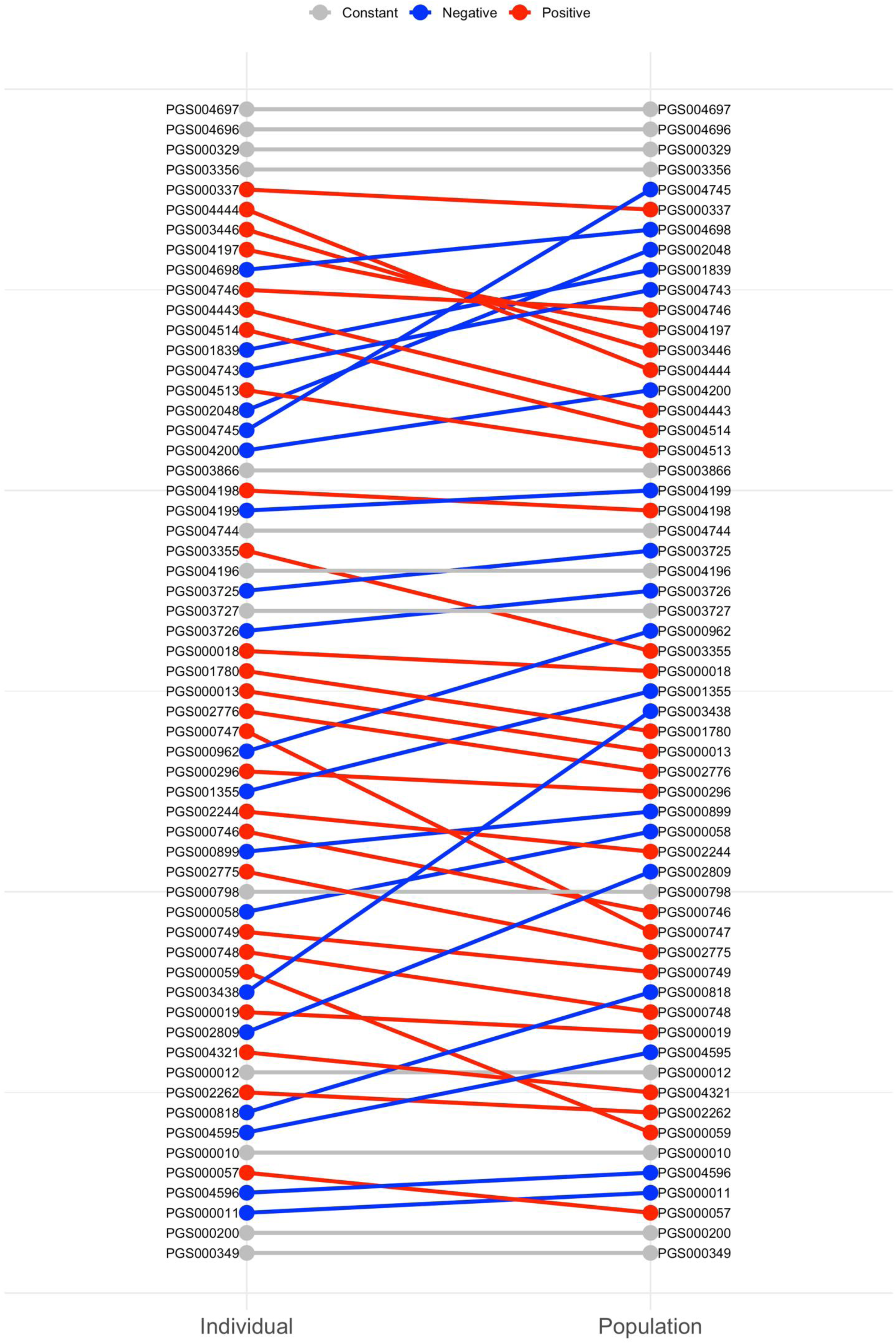
Rank transition of CAD PRSs at individual-level and population-level performance spaces. Red indicates positive slope: higher individual-level than population-level performance ranking. Blue indicates negative slope: higher population-level than individual-level performance ranking. Grey indicates constant slope and same performance ranking. Population-level performance was ranked by odds ratio per standard deviation, while individual-level performance was ranked by proportion of benchmark population (early onset, unexpected CAD) in the top decile of PRS distribution for all UKBB individuals.

### Characterizing Patients Missed by Current PRSs

Finally, we sought to understand why a subset of benchmark patients (early-onset, unexpected CAD) remains invisible to all of the 58 PRSs. We defined patients never captured in the top decile by any of the 58 PRSs (n=132) as those with “consistently low-PRS” and those frequently captured (identified by >20 of 58 scores, n=228) as those with “consistently high-PRS”. The primary distinction was a significantly healthier lipid profile in the consistently low-PRS group, who had higher LE8 blood lipid scores as well as lower Lp(a) concentrations, and lower PCE risk with fewer records of family heart disease history (**Table 3**). In contrast, the two groups showed no significant differences in demographics (sex, age, race, TDI), smoking and alcohol intake frequency, age of CAD onset, comorbidities (chronic kidney disease, rheumatoid arthritis, C reactive protein), CAD genetics (Familial Hypercholesterolemia and thrombophilia variant status) or overall CAD risk as assessed by the LE8 composite score (**Table 3**).

**Table 3.**
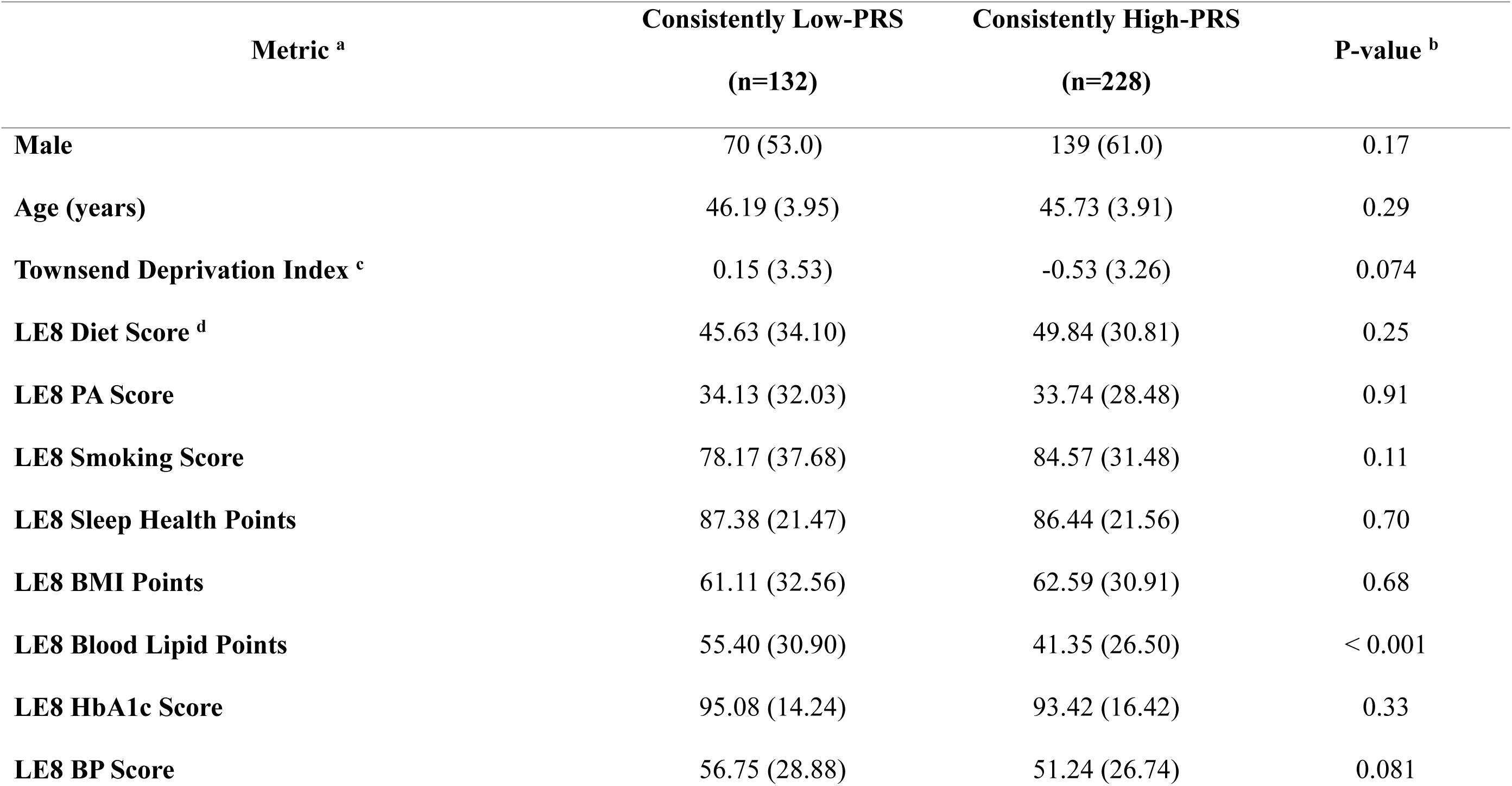
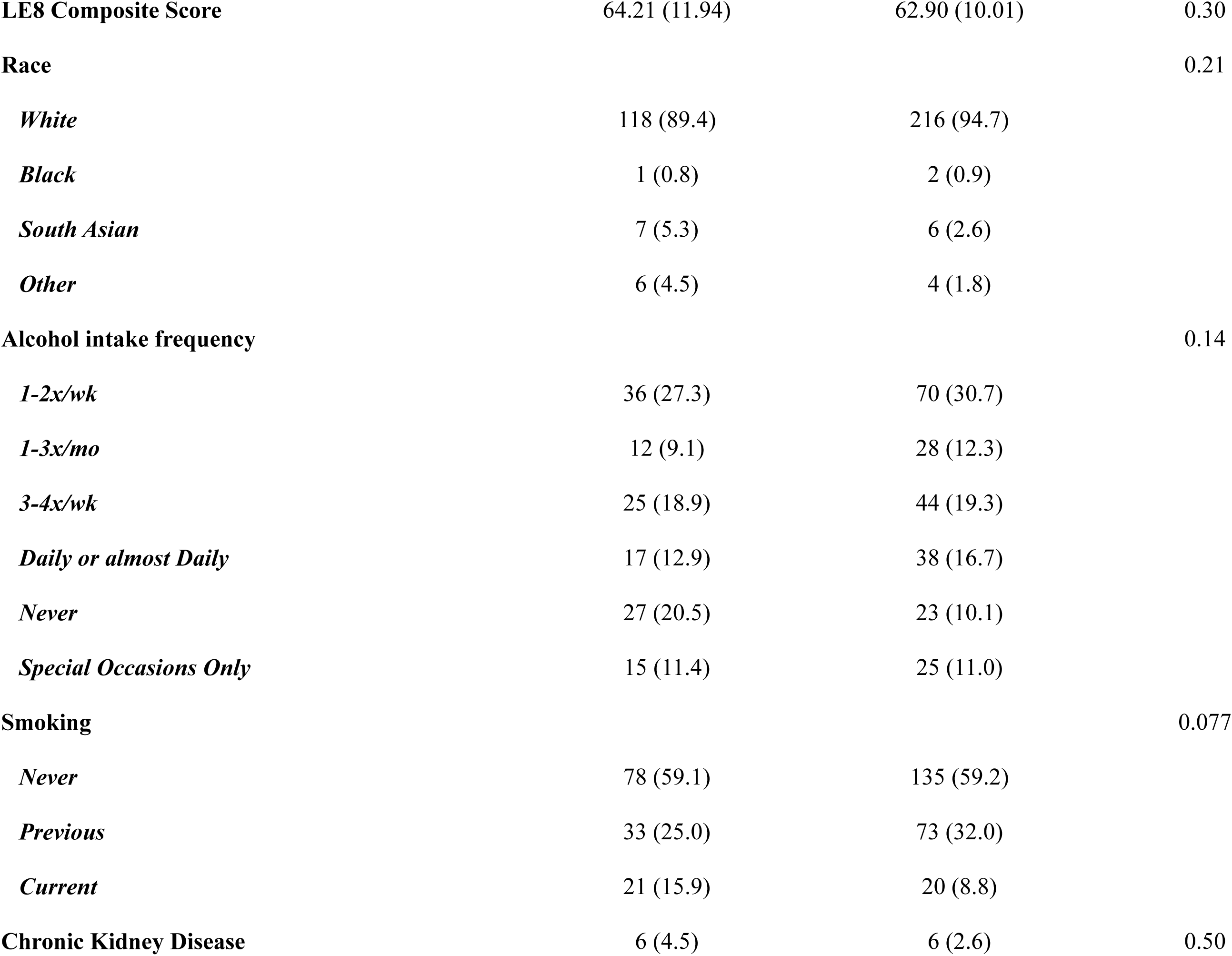
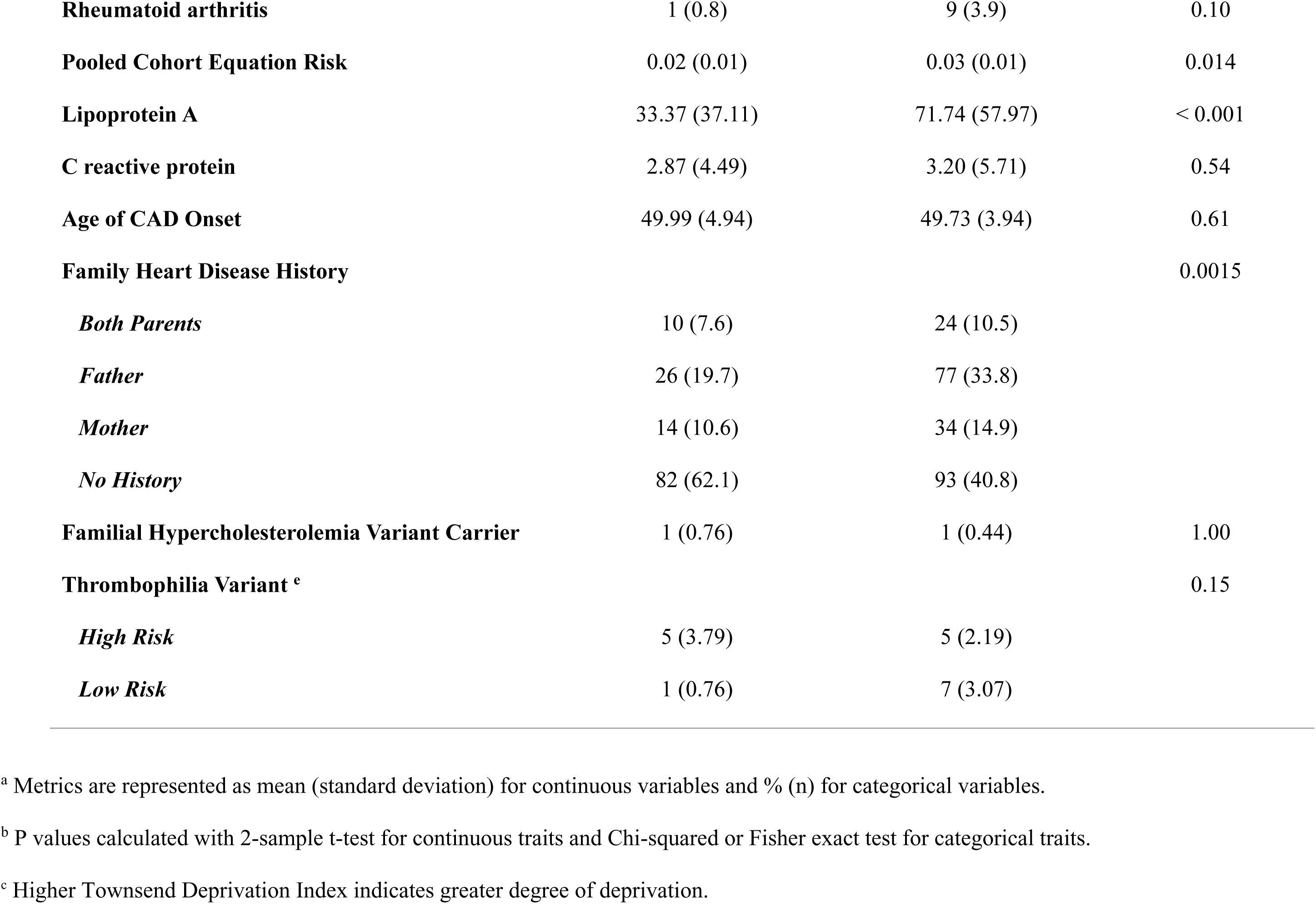
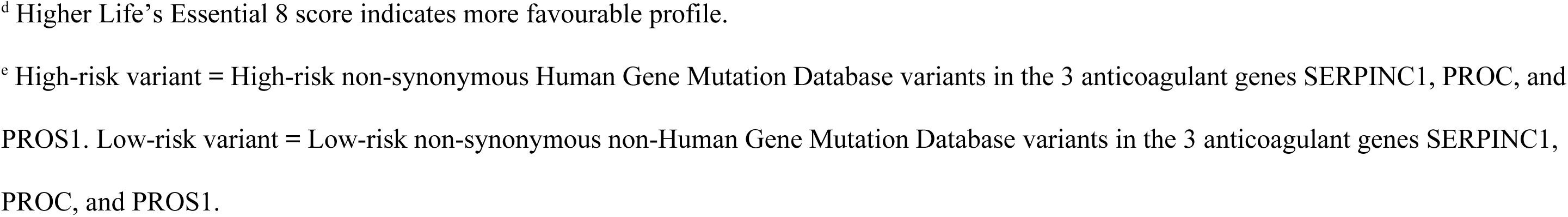
Individuals with early onset unexpected CAD stratified by how often they are captured by 58 CAD PRSs. The “Consistently Low-PRS” group is defined as individuals with early onset unexpected CAD who have never been identified as top 10% risk by any of the 58 PRS scores. The “Consistently High-PRS” group is defined as individuals with early onset unexpected CAD who have been identified as top 10% risk by more than 20 out of the 58 PRSs.

| **Metric <sup>a</sup>** | **Consistently Low-PRS**<br>**(n=132)** | **Consistently High-PRS**<br>**(n=228)** | **P-value <sup>b</sup>** |
| --- | --- | --- | --- |
| **Male** | 70 (53.0) | 139 (61.0) | 0.17 |
| **Age (years)** | 46.19 (3.95) | 45.73 (3.91) | 0.29 |
| **Townsend Deprivation Index <sup>c</sup>** | 0.15 (3.53) | -0.53 (3.26) | 0.074 |
| **LE8 Diet Score <sup>d</sup>** | 45.63 (34.10) | 49.84 (30.81) | 0.25 |
| **LE8 PA Score** | 34.13 (32.03) | 33.74 (28.48) | 0.91 |
| **LE8 Smoking Score** | 78.17 (37.68) | 84.57 (31.48) | 0.11 |
| **LE8 Sleep Health Points** | 87.38 (21.47) | 86.44 (21.56) | 0.70 |
| **LE8 BMI Points** | 61.11 (32.56) | 62.59 (30.91) | 0.68 |
| **LE8 Blood Lipid Points** | 55.40 (30.90) | 41.35 (26.50) | < 0.001 |
| **LE8 HbA1c Score** | 95.08 (14.24) | 93.42 (16.42) | 0.33 |
| **LE8 BP Score** | 56.75 (28.88) | 51.24 (26.74) | 0.081 |
| **LE8 Composite Score** | 64.21 (11.94) | 62.90 (10.01) | 0.30 |
| **Race** |  |  | 0.21 |
| *White* | 118 (89.4) | 216 (94.7) |  |
| *Black* | 1 (0.8) | 2 (0.9) |  |
| *South Asian* | 7 (5.3) | 6 (2.6) |  |
| *Other* | 6 (4.5) | 4 (1.8) |  |
| **Alcohol intake frequency** |  |  | 0.14 |
| *1-2x/wk* | 36 (27.3) | 70 (30.7) |  |
| *1-3x/mo* | 12 (9.1) | 28 (12.3) |  |
| *3-4x/wk* | 25 (18.9) | 44 (19.3) |  |
| *Daily or almost Daily* | 17 (12.9) | 38 (16.7) |  |
| *Never* | 27 (20.5) | 23 (10.1) |  |
| *Special Occasions Only* | 15 (11.4) | 25 (11.0) |  |
| **Smoking** |  |  | 0.077 |
| *Never* | 78 (59.1) | 135 (59.2) |  |
| *Previous* | 33 (25.0) | 73 (32.0) |  |
| *Current* | 21 (15.9) | 20 (8.8) |  |
| **Chronic Kidney Disease** | 6 (4.5) | 6 (2.6) | 0.50 |
| **Rheumatoid arthritis** | 1 (0.8) | 9 (3.9) | 0.10 |
| **Pooled Cohort Equation Risk** | 0.02 (0.01) | 0.03 (0.01) | 0.014 |
| **Lipoprotein A** | 33.37 (37.11) | 71.74 (57.97) | < 0.001 |
| **C reactive protein** | 2.87 (4.49) | 3.20 (5.71) | 0.54 |
| **Age of CAD Onset** | 49.99 (4.94) | 49.73 (3.94) | 0.61 |
| **Family Heart Disease History** |  |  | 0.0015 |
| *Both Parents* | 10 (7.6) | 24 (10.5) |  |
| *Father* | 26 (19.7) | 77 (33.8) |  |
| *Mother* | 14 (10.6) | 34 (14.9) |  |
| *No History* | 82 (62.1) | 93 (40.8) |  |
| **Familial Hypercholesterolemia Variant Carrier** | 1 (0.76) | 1 (0.44) | 1.00 |
| **Thrombophilia Variant <sup>c</sup>** |  |  | 0.15 |
| *High Risk* | 5 (3.79) | 5 (2.19) |  |
| *Low Risk* | 1 (0.76) | 7 (3.07) |  |

<sup>a</sup> Metrics are represented as mean (standard deviation) for continuous variables and % (n) for categorical variables.
<sup>b</sup> P values calculated with 2-sample t-test for continuous traits and Chi-squared or Fisher exact test for categorical traits.
<sup>c</sup> Higher Townsend Deprivation Index indicates greater degree of deprivation.
<sup>d</sup> Higher Life's Essential 8 score indicates more favourable profile.
<sup>e</sup> High-risk variant = High-risk non-synonymous Human Gene Mutation Database variants in the 3 anticoagulant genes SERPINC1, PROC, and PROS1. Low-risk variant = Low-risk non-synonymous non-Human Gene Mutation Database variants in the 3 anticoagulant genes SERPINC1, PROC, and PROS1.

### Sensitivity Analysis: Investigating Variations in Benchmark Cohort Definitions

To validate our findings under the most rigorous conditions possible, we replicated our study with a series of sensitivity analyses (**Table 4**). Baseline characteristics of benchmark cohorts from sensitivity analysis 1-4 were summarized in **Supplementary Table 3**. Sensitivity analysis 1-3 varied the definitions of the benchmark cohort used for our individual-level metric, sharing the same population metric model as that of the primary analysis (**Supplementary Table 4**). Sensitivity analysis 4 used a stricter definition of “unexpected CAD” from published criteria^7^. This definition excluded individuals with any history of hypertension, diabetes, dyslipidemia, or smoking, as well as those taking relevant medications (**Supplementary Table 7**). In sensitivity analysis 4, we also varied the population-level metric model and utilized Cox proportional hazards models to estimate the HR of CAD per SD increase in PRS (**Supplementary Table 8**).

**Table 4.**
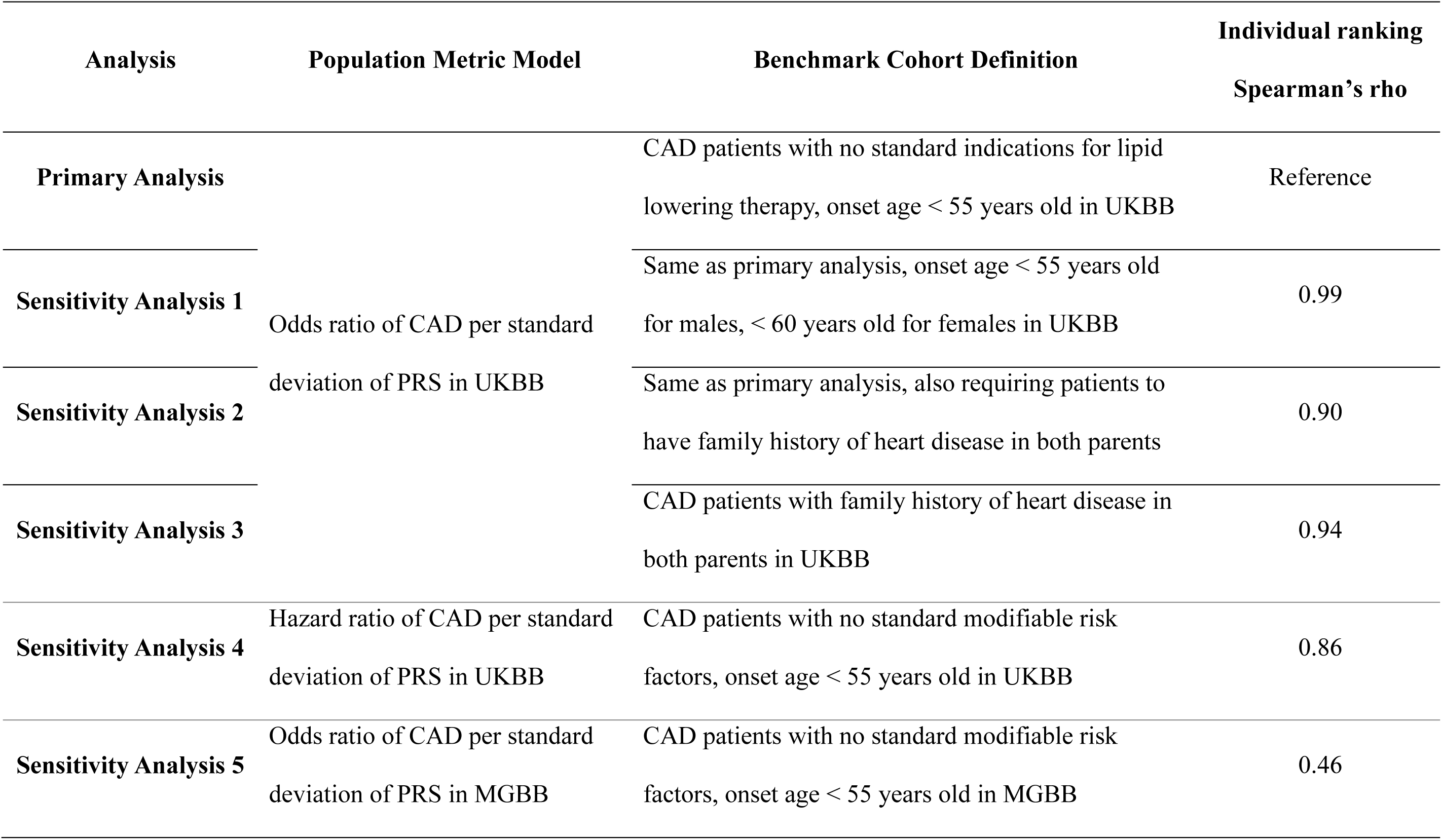
Population metric model and benchmark cohort definition in the primary analysis and sensitivity analysis 1-5. Spearman’s rho was calculated to compare individual-level ranking of each sensitivity analysis with the primary analysis.

Finally, we replicated the cohort definition of sensitivity analysis 4 in the Massachusetts General Brigham Biobank (MGBB), acknowledging limitations due to the overall European-dominant population profile of UKBB and the potential of overfitting when evaluating PRS performances in the same cohort that supplied genome-wide analysis study (GWAS) results for their training. Population-level metric for the MGBB replication follows an OR model similar to that of the primary analysis (**Supplementary Table 10**).

Despite these variations in benchmark cohort definitions, the results closely mirrored the primary analysis as suggested by Spearman’s rho (**Table 4**), while the modest but positive correlation between individual-level PRS ranking of our primary analysis and our MGBB replication (sensitivity analysis 5) was expected due to differences in nature of data, curation processes and population ancestries.

Multi-ancestry scores again demonstrated higher consistency across ancestry subgroups (sensitivity analysis 1-4: British White and non-British White; sensitivity analysis 5: non-Hispanic White and other ancestry) for both individual-level and population-level metrics (**Supplementary Figures 3 & 5**). Wider CIs of percentage captures in subgroup analysis of sensitivity analysis 2, 4 and 5 were the result of smaller benchmark cohorts used for individual-level evaluations. While sensitivity analysis 2 and 4 operationalized stricter definition of high genetic risk CAD patients leading to reduced sizes of benchmark cohort, results from sensitivity analysis 5 reflected difficulties in validating CAD patients in EHR data, leading to similarly wide CIs in population-level ORs of the non-white ancestry subgroup, not observed in analysis using UKBB (**Supplementary Figure 5**). On the other hand, sensitivity analysis 1 and 3, along with the primary analysis, remained of power to detect differences in benchmark cohort capture rates in ancestry- and sex-based subgroups (**Supplementary Figure 3 & 4**).

The top PRS identified by our individual-level metric remained an effective tool for identifying the high genetic risk benchmark cohort across differences in definitions of such cohort, with significant improvement in benchmark cohort identification compared with single clinical risk factors Lp(a), CRP and BMI (**Supplementary Figure 6**). The more similar performances comparing top PRS and the combinatory metric of PRS + Lp(a), as well as the overlapping performances comparing top PRS and the combinatory metric of PRS + PCE, underscored the importance of evaluating individual-level performances of PRSs for selection of the best towards clinical application. In all cases, the PRS used for deriving the combinatory metrics were the top-performing score from the respective individual ranking of each sensitivity analysis. This analysis was not replicated in sensitivity analysis 5 with MGBB data, due to difficulty in ascertaining clinical risk factors at time of diagnosis and the potential of biased results.

Albeit with slightly different ranking and top performing scores, the conclusion on Bayesian methods^23^, P+T^26^ and penalized regression scores were consistent with primary analysis. Specifically, top-performed scores, for both population-level and individual-level metrics, remained genome-wide scores using Bayesian approaches; scores involving variant selection by P+T or penalized regression have good individual-level performance with slight drop in population-level performance (**Supplementary Figure 7, Supplementary Table 4, 8 & 10**). Worth noticing, while the individual-level ranking from MGBB replication was only at a Spearman’s rho of 0.46 with our primary analysis with UKBB, Bayesian method scores occupied top 10 of the MGBB individual ranking, adding weight to our conclusions. Although not top performing in the primary analysis, multi-trait and multi-method scores reached top 10 in sensitivity analysis using family history in benchmark cohort definition, and sensitivity analysis using MGBB. PGS004697^17^ (PRS-CSx^18^, multi-ancestry) remained the top-performing score on both population- and individual-level throughout the primary analysis and sensitivity analysis 1-3 (**Supplementary Figure 7**).

Finally, the profile of the “invisible” patient persisted. Consistently low-PRS benchmark patients (never captured by any score) were distinguished from consistently high-PRS benchmark patients (frequently captured patients) most significantly by healthier lipid profiles underlying lower PCE risk and fewer records of family heart disease history (**Supplementary Table 5, 6 & 9**). Analysis by capture frequency was not conducted for sensitivity analysis 2 due to insufficient benchmark cohort size (n=109) or sensitivity analysis 5 due to difficulty ascertaining baseline clinical biomarker levels in MGBB linked EHR.

## Discussion

In this study, we developed and applied a benchmark framework to address the well-documented disconnection between the population-level performance of PRSs and their utility for individual-level risk prediction^1,5,6^. While prior studies have established the conceptual problem of individual-level PRS variability, our contribution is the proposal and application of an operational, clinically grounded benchmark cohort that enables direct, head-to-head evaluation of individual-level PRS performance in the precise scenario where a genetic risk tool would be most valuable. To our knowledge, this work is the first to systematically evaluate a large panel of published CAD PRSs against a clinically-defined “ground truth” cohort of early-onset, unexpected CAD patients—individuals whose disease is most plausibly driven by high inherited risk theoretically identifiable by PRS. Our approach progresses PRS validation from relative, population-based comparisons to an absolute-scale assessment of clinical accuracy. We demonstrated that this framework not only revealed profound variability in individual-level performances (benchmark capture rates from 10.8% to 33.1%) that is obscured by traditional metrics, but also uncovered key biological limitations of current scores. With consistent results from both our primary analysis and a more stringent definition of unexpected CAD in the sensitivity analysis, we showed that this benchmark paradigm provides a robust, generalizable blueprint for evaluating one essential dimension of PRS performance, individual-level risk identification, which is a necessary step toward establishing the clinical readiness of PRSs across complex diseases.

Our findings on development methodology provided a potential explanation for the observed performances. In this specific cohort, we found that Bayesian-based approaches^23^ (e.g., LDpred2^28^, PRS-CSx^18^) were hallmarks of robust, concordant performance: the Bayesian-derived scores that ranked in the individual-level top 10 also all ranked in the population-level top 10. This suggests these methods may be effective at capturing risk across both evaluation paradigms. Another group of scores that were top-performing at the individual-levels was developed with P+T or penalized regression methods. However, this group’s performance was less prominent on the population level compared to the Bayesian scores.

Multi-trait and multi-method scores were generally not among the top performed scores based on individual-level metrics. This was particularly evident when comparing scores (PGS004443, PGS004444, PGS004513, PGS004514)^14^ derived from the same data and same variants, where LDpred2-based^28^ scores performed better at individual-level risk prediction. This suggested that incorporating correlated traits or methods might offer potentially limited value for identifying specific, high-risk individuals.

Beyond methodological differences, our benchmark framework also exposed a critical biological limitation of current CAD PRSs. Our analysis of the benchmark cohort revealed that individuals with consistently low PRS never captured by the top decile of any of the 58 scores had significantly healthier lipid profiles and lower Lp(a) concentrations than those with consistently high PRS frequently captured. This finding strongly implied that the current generation of PRSs might be heavily weighted towards variants in lipid-mediated pathways^29,30^, likely due to the high polygenicity and strong genetic signals of lipids. Consequently, they systematically failed to identify a distinct subgroup of high-risk patients whose disease is likely driven by non-lipid genetic etiologies. Several candidate mechanisms may underlie this subgroup’s risk. Inflammatory pathways represent one plausible axis, given that multiple CAD GWAS loci map to genes involved in cytokine signaling (e.g., IL6R, SH2B3) and leukocyte recruitment^20^, and that anti-inflammatory therapies reduce cardiovascular events independently of lipid lowering^31^. Additionally, clonal hematopoiesis of indeterminate potential (CHIP), driven by somatic mutations in *DNMT3A*, *TET2*, and *JAK2*, accelerates atherosclerosis through inflammasome-mediated pathways entirely independent of lipid metabolism^32^. Future PRS development efforts that specifically incorporate or upweight variants in these non-lipid pathways may improve identification of the high-risk patients currently missed by existing scores.

Despite this clear bias, the clinical potential of PRS remains evident. The best-performing score PGS004697^17^ captured 33.1% of our benchmark cohort, yet this was more than 2-fold higher than the proportion captured by established biomarkers like Lp(a), CRP, or BMI (∼12%). This demonstrated the unique value of PRS in identifying high-risk individuals who would have been missed by standard clinical assessments, especially when combining the top-performing PRS at capturing high genetic risk benchmark cohort CAD patients with existing risk estimators like PCE. However, to realize the full potential of genomic medicine, our findings highlighted a need for future score development to specifically target variants that confer CAD risk through non-lipid pathways. This is essential for accurately identifying all patients with high inherited risk.

Our benchmark framework is timely in the context of accelerating efforts to translate PRS into clinical practice. Recent studies have demonstrated the feasibility of returning combined monogenic and polygenic risk results to patients in preventive genomics clinics, with meaningful impacts on clinical management including statin initiation and coronary imaging^33^. Moreover, integrating Lp(a) with CAD PRS has been shown to modestly improve risk prediction^34^, and there is growing recognition that the most effective risk stratification will come from models that incorporate both genetic and clinical data in combination. From a clinical standpoint, a high PRS now serves as a risk enhancer alongside family history, Lp(a), and chronic inflammatory conditions under the current guideline^35^, and prospective trials such as PROACT1 (NCT05819814) are now evaluating whether disclosing high polygenic risk leads to improved clinical outcomes. As PRS moves closer to clinical deployment in these settings, frameworks like ours that provide absolute-scale, clinically interpretable validation of individual-level PRS performance will be essential for ensuring that the scores used in practice can reliably identify the patients who stand to benefit most from early intervention.

Our conclusions were robust against variations in benchmark cohort definitions, population-level metric models and dataset used, as demonstrated by our sensitivity analyses. We utilized a series of definitions for our benchmark cohort of high genetic risk CAD patients, leveraging requirements on sex-specific onset age, absence of standard indicators for lipid-lowering medications, absence of standard modifiable risk factors, and family history of heart disease in both parents. We also varied our population-level metric model using both OR and HR per SD of PRS. Most importantly, we successfully replicated our findings in MGBB data. This replication addressed limitations of our UKBB-based analysis due to potential of overfitting that inflates effect estimates and the mostly European^36^ composition of UKBB populations. We also evaluated ranking changes in non-UKBB-derived scores only (**Supplementary Table 11**).

However, this study has several limitations. Firstly, several reasons may affect benchmark cohort curation results. For the primary analysis, low 10-year risk and severe hypercholesterolemia definition leveraged the 2018 AHA guidelines^8,9^, but updates may be available. On the other hand, sensitivity analyses 2 and 3 relied on data fields 20107 and 20110 from UKBB, which were self-reported family history of heart disease, arguably a wider definition than CAD central to our analysis. In sensitivity analysis 4, not all criteria from Figtree Gemma et al.^7^ were used in the curation of the unexpected CAD population; for example, fasting and oral glucose tolerance test 2-h blood glucose levels were not available in UKBB for enough participants, and we excluded them in benchmark cohort curation to avoid losing too many patients due to missingness.

Second, although our replication in MGBB showed agreeing results, patient information in this dataset was available through linked retrospective EHR, posing challenges in identification and validation of the benchmark cohort, especially compared with data quality from prospective cohorts like UKBB where patient records of diagnosis dates and clinical measurements were available from the time of database enrollment to CAD. Specifically, while we carried out strict data processing steps, we could not ensure that all measurements were made at the very date of CAD diagnosis due to data availability limitations. Moreover, diabetes mellitus medication status was not used in curation due to difficulty separating patients taking such medications for diabetes and obesity. Nonetheless, we restricted biomarker measurement date to that closest to CAD diagnosis and before risk factor manifestation to avoid reverse causation and misclassification from using post-medication data.

Our work provided a new, absolute-scale framework for validating PRSs, confirming a critical disconnect between population-level metrics and individual-level accuracy^1,5,6^. We demonstrated that this variability was not random but could be driven by development methodology and by a biological blind spot. With the top-performing PRSs already showed clear clinical utility, outperforming biomarkers like Lp(a) by more than 2-fold, our benchmark approach highlighted a need to correctly target and weight multiple disease pathways. This framework is essential for guiding the development of the next generation of PRSs that can reliably identify all high-risk individuals. Moreover, our results carry a practical implication for clinicians and researchers selecting among available PRSs: when multiple scores show similar population-level performance (e.g., comparable AUC or OR per SD), our framework demonstrates that they can differ substantially in their ability to identify individual patients at the highest genetic risk. Validation against clinically defined extreme phenotypes, such as the benchmark cohort used here, should therefore be considered an essential step before deploying any PRS in patient care.

## Methods

### Study population

The UK Biobank (UKBB) is a prospective, observational, population-based cohort of the United Kingdom, enrolling >500,000 adult residents aged 40-69 years at the time of recruitment who provided information on medical history, medication use, lifestyle factors at baseline, underwent physical assessment, laboratory analysis and genotyping, and were followed for development of incident diagnoses confirmed by linkage to national health records^37^. The secondary use of data for the present analysis was approved by the Massachusetts General Hospital Institutional Review Board (protocol 2021P002228) and facilitated through UK Biobank Application 7089.

### Curation of unexpected CAD population

The definition of CAD was based on a combination of ICD9, ICD10 and OPCS-4 (Office of Population Censuses and Surveys Classification of Interventions and Procedures version 4) codes as well as self-reported illness and doctor diagnosis (**Supplementary Table 1**). The criteria for unexpected CAD included: low 10-year risk (Pooled Cohort Equation [PCE] risk < 0.05^8^), absence of diabetes (hemoglobin A1c [HbA1c] < 6.5%, no diabetes mellitus medication) and no severe hypercholesterolemia (low-density lipoprotein cholesterol [LDL-C] < 4.9 mmol/L^9^, no lipid-lowering medication). From the full UKBB cohort, our study population included participants who had complete records of above criteria to define the benchmark cohort of early onset unexpected CAD. After applying these criteria, the final analytical cohort comprised 41,397 participants. Expected CAD was defined as those having CAD and complete data on definitive criteria, but not qualified as unexpected CAD of strong genetic predisposition. Early onset age cutoff is 55 years old.

### PRS calculation

The variant effect sizes for PRS of CAD were downloaded from Polygenic Score Catalog^38^ (https://www.pgscatalog.org/). Searches were conducted using the term “coronary artery disease” as of June 20, 2024. All PRS calculations were performed using the PLINK2^39^ software via the --score function. We adjusted all polygenic scores for enrollment age, sex, genotyping array, and the first four principal components of genetic ancestry. Subsequently, scores were standardized to have a mean of zero and a standard deviation of one, ensuring uniformity across evaluations. All polygenic scores were residualized for the first four principal components of genetic ancestry and then scaled to a mean of 0 and a standard deviation of 1.

To assess the correlation among PRS, we utilized the corrplot function in R. Four scores were excluded from the analysis (PGS000116^40^, PGS004899^41^, PGS004888^42^, PGS004237^43^) due to their negative correlations with the majority of other scores. Following these exclusions, 58 CAD scores remained for further analysis.

### Identification of early-onset unexpected CAD population by 58 PRSs

Out of 451,580 UKBB individuals with calculated scores, the proportion of early onset unexpected CAD identified as having top 10% PRS was calculated for each of the 58 PRSs with 95% CIs obtained by bootstrapping. This application of general CAD PRSs to an early-onset subgroup is supported by evidence that PRS effect sizes for CAD are consistently larger in younger individuals than in older ones^43,44^, suggesting that common risk variants are concentrated, rather than absent, in cases unexplained by conventional risk factors.

Individual-level performance ranking of PRS was based on proportion identification in all UKBB population regardless of ancestry, but we also evaluated proportion in British White and non-British White populations to understand a score’s performance across ancestries. Sex-stratified capture analysis was carried out as well. We also used top 10% BMI, Lp(a) or CRP instead of top 10% PRS to compare the ability of PRS vs. single risk factor representing obesity, lipid profile and inflammation in capturing benchmark high genetic risk CAD patients. To take into account combinational metrics closer to those used in clinical practice, we used the top-performing PRS and combined with standardized Lp(a) (mean of 0 and standard deviation of 1) and PCE, respectively. McNemar’s test was performed to evaluate if the proportion of benchmark cohort identified by each variable was statistically significantly different than that identified by the top PRS.

### Population-level performance metric of 58 PRSs

For the population-level evaluation, cases included all UKBB participants with CAD (encompassing both expected and unexpected CAD) and controls included all UKBB participants without CAD. This is intentional: the population-level metric is the conventional approach to evaluating PRS performance (OR per SD), and it is deliberately contrasted against our novel individual-level benchmark metric. The benchmark cohort (early onset, unexpected CAD) was used exclusively for the individual-level evaluation. We used the odds ratio (OR) of CAD per standard deviation increase in PRS as the population-level performance metric. The logistic regression model was adjusted for age, sex, and the first four principal components of genetic ancestry. Same as visualization of individual performance metric, population-level performance ranking of PRS was based on OR per standard deviation in all UKBB population regardless of ancestry, but we also evaluated OR per standard deviation in British White and non-British White populations to understand a score’s performance across ancestries.

### Ranking comparison between population- and individual-level performance

Slope plots were used to compare population- and individual-level performance ranking of scores in all UKBB subjects regardless of race. PRSs were further categorized into three non-mutually exclusive groups: multi-ancestry, multi-traits and multi-methods scores. Multi-ancestry scores used multi-ancestry source of variant association or score development populations evident from the PGS catalog^11,12^. Multi-trait scores were identified by “incorporation of genetically correlated traits/CAD-related traits/risk factors” or similar wordings in their source publications. Multi-method scores referred to those developed by leveraging multiple scores of the same trait as made clear in source publications.

### Individuals with early onset, unexpected CAD stratified by frequency of being identified by 58 CAD PRSs

Acknowledging that reasons behind differences in rankings of individual-level performance of the 58 PRSs went beyond score development methods, we compared a series of characteristics, including demographics (sex, TDI, age, race), lifestyle factors (diet, physical activities, smoking, sleep, alcohol intake), CAD-related clinical measurements (age of onset, BMI, blood lipid, HbA1c, blood pressure, chronic kidney disease, rheumatoid arthritis, pooled cohort equation risk, LE8 composite score, C reactive protein) and CAD genetics (family history of heart disease, Familial Hypercholesterolemia carrier status, thrombophilia variant status) between individuals frequently identified in the top 10% of the UKBB by the 58 scores, and those never identified in the top 10% out of UKBB by any score.

## Sensitivity Analysis

### Study population

Sensitivity analysis 1-4 utilized the same study population as the primary analysis. In sensitivity analysis 5, we used data from the Massachusetts General Brigham Biobank (MGBB). MGBB is an integrated research initiative based in Boston, Massachusetts, collecting biological samples and health data from consenting individuals at Massachusetts General Hospital, Brigham and Women’s Hospital, and local healthcare sites within the MGB network^45,46^. The secondary use of data for the present analysis was approved by the Massachusetts General Hospital Institutional Review Board (protocol 2021P002228).

### Curation of unexpected CAD population

Sensitivity analysis 1-3 used alternative definitions of the benchmark cohort for our individual-level metric. Sensitivity analysis 1 imposed sex-differential early onset age cutoff (55 years old for males and 60 years old for females) to account for later CAD manifestation in females.

Sensitivity analysis 2 and 3 explored further definitions of high genetic risk CAD patients using UKBB data fields 20107 and 20110 as source of heart disease family histories. In sensitivity analysis 2, we selected those within our primary analysis benchmark cohort that also had family history of heart disease in both parents, while in sensitivity analysis 3, we used family history of heart disease in both parents as the only definition of high genetic risk CAD.

The criteria for a more stringent unexpected CAD definition using the absence of standard modifiable risk factors in sensitivity analysis 4 were described by Figtree Gemma et al^7^. Briefly, the standard modifiable risk factors evaluated at baseline (UKBB enrollment) included smoking, hypertension, diabetes mellitus, and dyslipidemia (**Supplementary Table 4**). To ensure a strict temporal order and avoid reverse causation from classifying post-medication measurements, we implemented a follow-up time check and excluded those with CAD outcomes developed less than 6 months before diagnosis of any risk factor. Early onset age cutoff was still set at 55 years old. A cohort of CAD patients with standard modifiable risk factors was also curated including those that were not unexpected CAD with no missing value in any of the above-mentioned criteria.

Finally, we replicated the cohort definition of sensitivity analysis 4 (**Supplementary Table 4**) in MGBB out of 65,183 individuals with available genotype and phenotype data. We preserved the strict temporal order in patient profile evaluation by searching for linked EHR records closest to CAD diagnosis date and before risk factor diagnosis dates of each patient, to avoid reverse causation and misclassification bias from evaluating post-medication biomarkers. Diabetes medication status was not curated in the MGBB-linked EHR, since data limits the potential of distinguishing between patients taking glucose-lowering medications for diabetes and obesity. A main difference from data curation using the prospective UKBB is that in MGBB linked EHR, we kept patients with missingness, assuming they did not have risk factors if there were no records against such assumption. Corresponding to the ancestry classification of UKBB, the benchmark cohort was divided into non-Hispanic White and other ancestry subgroups.

### PRS calculation

Sensitivity analysis 1-4 shared the same calculated PRS as the primary analysis and methods were described in previous section. PRS calculation in MGBB used the same pipeline as that in UKBB.

### Identification of early-onset unexpected CAD population by 58 PRSs

Sensitivity analysis 1-4 shared the same method of obtaining individual-level performance metric of PRSs as the primary analysis, with only changes made to the definitions of early-onset unexpected CAD benchmark cohorts. Sensitivity analysis 5 used the same method applied to a different data source: out of 49,744 MGBB individuals with calculated scores, the proportion of early onset unexpected CAD identified as having top 10% PRS was calculated for each of the 58 PRSs with 95% CIs obtained by bootstrapping. We presented percentages of benchmark cohort captured by ancestry (UKBB: British White and non-British White; MGBB: non-Hispanic White and other) and sex subgroups.

Similar to the primary analysis, top PRS was compared with 3 single clinical biomarkers (BMI, Lp(a), CRP) and 2 combinational metrics (PRS + standardized Lp(a), PRS + PCE) and McNemar’s test was performed to obtain p-values. For all sensitivity analyses, the PRS used for deriving the combinatory metrics were the top-performing score from the respective individual ranking of each sensitivity analysis (sensitivity analysis 1-3: PGS004697^17^; sensitivity analysis 4: PGS004444^14^). This comparison was not replicated in sensitivity analysis 5 using MGBB data due to difficulty in ascertaining clinical biomarkers at time of CAD diagnosis in EHR data.

### Population-level performance metric of 58 PRSs

Sensitivity analysis 1-3 shared the same population metric model as that of the primary analysis using odds ratio per standard deviation of PRS as effect estimate. In sensitivity analysis 4, the population-level evaluation included all UKBB participants free of both prevalent CAD and prevalent risk factors (hypertension, diabetes, hypercholesterolemia) at baseline. Follow-up began at enrollment. We used hazard ratio (HR) of CAD per standard deviation increase in PRS as the population-level performance metric. To ensure positive follow-up time, we excluded those with prevalent risk factors or CAD diagnosis. Individuals were censored at the time of developing any defining risk factor (diabetes, hypercholesterolemia, or hypertension), ensuring that the Cox regression evaluated only person-time free of conventional modifiable risk factors. In sensitivity analysis 5, population-level metric for the MGBB replication followed an odds ratio (OR) model similar to that of the primary analysis, switching the 451,580 UKBB individuals with calculated scores to 49,744 MGBB individuals. All results were presented in the same way as the primary analysis, stratifying by ancestry.

### Ranking comparison between population- and individual-level performance

Slope plots were used to compare population- and individual-level performance ranking of scores in all UKBB (sensitivity analysis 1-4) or MGBB (sensitivity analysis 5) subjects regardless of race. Individual-level benchmark definitions and corresponding population-level metric models were detailed in previous method sections, as well as **Table 4**.

### Individuals with early onset, unexpected CAD stratified by frequency of being identified by 58 CAD PRSs

Similar to the primary analysis, we compared a series of characteristics, including demographics, lifestyle factors, CAD-related clinical measurements and CAD genetics between individuals frequently (by > 20 of the 58 scores) identified in the top 10% out of UKBB by the 58 scores, and those never identified in the top 10% out of UKBB by any score. Analysis by capture frequency was not conducted for sensitivity analysis 2 due to insufficient benchmark cohort size (n=109) and sensitivity analysis 5 due to difficulty ascertaining baseline clinical biomarker levels in MGBB-linked EHR. Variables central to the definition of the benchmark cohort were not compared between groups since all individuals, by definition, had the same status, for example, family history of heart disease in sensitivity analysis 3, and smoking and Familial Hypercholesterolemia carrier status in sensitivity analysis 4.

## Data availability

For primary analysis and sensitivity analysis 1-4, data supporting the results of the present study are available from the UKB (https://www.ukbiobank.ac.uk/enable-your-research/apply-for-access) to bona fide researchers with institutional review board and UKB approval. These analyses were performed using the UKB resource under application no. 7089. The secondary use of these data was approved by the Mass General Brigham institutional review board. Sensitivity analysis 5 used data from Massachusetts General Brigham biobank and linked EHR. The secondary use of data for the present analysis was approved by the Massachusetts General Hospital Institutional Review Board (protocol 2021P002228).

## Code availability

All analyses were performed using R and PLINK, as detailed in Methods. No novel computational methods or custom codes were developed that were essential to the paper’s conclusions. All analytical scripts are openly available via GitHub at https://github.com/zhiyulab/CAD_PRS.git.

## Data Availability

All data produced in the present study are available upon reasonable request to the authors

## Acknowledgements

All UKB analyses were performed under application no. 7089. Dr. Yu is supported by R00(HG012956) from National Human Genome Research Institute and Early Career Investigator Award (18CVD04) from Foundation Leducq Clonal Hematopoiesis and Atherosclerosis Network. Dr. Fahed is supported by grants K08HL161448 and R01HL164629 from the National Heart, Lung, and Blood Institute. Dr. Sui is supported by the TOPMed fellowship from the National Heart Lung and Blood Institute.

## Author Information

These authors contributed equally: Shengxin Liang, Min Seo Kim. These authors jointly supervised this work: Akl C. Fahed, Zhi Yu.

## Contributions

M.S.K., Y.S., P.N., N.C., A.C.F. and Z.Y. designed the study. S.L. and Y.S. performed data analysis. K.P. contributed data on MGBB CAD phenotypes. Y.L. contributed data on Life’s Essential 8 scores. S.L., M.S.K., A.C.F., Z.Y. interpreted the study findings. S.L., Y.S., M.S.K and Z.Y. drafted the paper. Y.T. contributed codes for data analysis. All authors reviewed the paper and provided critical feedback.

## Ethics Declarations

Inclusion and ethics standards have been reviewed where applicable.

