## Supplemental Table 1-11 for "Evaluating Individual Level Performance of Polygenic Risk Scores Using Early Onset High Genetic Risk Coronary Artery Disease as a Benchmark"

**Supplementary Table 1. Codes of UK Biobank (UKBB) data fields used to define coronary artery disease (CAD).**

| Field | Description | Coding |
|---|---|---|
| **20002** | Non-cancer illness code, self-reported | 1075 |
| **20004** | Operation code | 1070,1095,1523 |
| **6150** | Vascular/heart problems diagnosed by doctor | 1 |
| **41202** | Diagnosis – main ICD10 | I21,I21.0,I21.1,I21.2,I21.3,I21.4,I21.9,I22,I22.0,I22.1,I22.8,I22.9,I23,I23.0,I23.1,I23.2,I23.3,I23.4,I23.5,I23.6, I23.8,I24,I24.0,I24.1,I24.8,I24.9,I25.1,I25.2,I25.5,I25.6,I25.8,I25.9 |
| **41204** | Diagnosis – Secondary ICD10 | I21,I21.0,I21.1,I21.2,I21.3,I21.4,I21.9,I22,I22.0,I22.1,I22.8,I22.9,I23,I23.0,I23.1,I23.2,I23.3,I23.4,I23.5,I23.6, I23.8,I24,I24.0,I24.1,I24.8,I24.9,I25.1,I25.2,I25.5,I25.6,I25.8,I25.9 |
| **40001** | Underlying (primary) cause of death: ICD10 | I21,I21.0,I21.1,I21.2,I21.3,I21.4,I21.9,I22,I22.0,I22.1,I22.8,I22.9,I23,I23.0,I23.1,I23.2,I23.3,I23.4,I23.5,I23.6, I23.8,I24,I24.0,I24.1,I24.8,I24.9,I25.1,I25.2,I25.5,I25.6,I25.8,I25.9 |
| **40002** | Contributory (secondary) causes of death: ICD10 | I21,I21.0,I21.1,I21.2,I21.3,I21.4,I21.9,I22,I22.0,I22.1,I22.8,I22.9,I23,I23.0,I23.1,I23.2,I23.3,I23.4,I23.5,I23.6, I23.8,I24,I24.0,I24.1,I24.8,I24.9,I25.1,I25.2,I25.5,I25.6,I25.8,I25.9 |
| **41200** | Operative procedures – main OPCS4 | K40,K40.1,K40.2,K40.3,K40.4,K40.8,K40.9,K41,K41.1,K41.2,K41.3,K41.4,K41.8,K41.9,K42,K42.1,K42.2, K42.3,K42.4,K42.8,K42.9,K43,K43.1,K43.2,K43.3,K43.4,K43.8,K43.9,K44,K44.1,K44.2,K44.8,K44.9,K45.1, |

| | | |
|---|---|---|
| | | K45.2,K45.3,K45.4,K45.5,K45.6,K45.8, K45.9,K46,K46.1,K46.2,K46.3,K46.4,K46.5,K46.8,K46.9,K49.1, K49.2,K49.3,K49.4,K49.8,K49.9,K50.1,K50.2,K50.4,K75.1,K75.2, K75.3,K75.4,K75.8,K75.9 |
| **41210** | Operative procedures – Secondary OPCS4 | K40,K40.1,K40.2,K40.3,K40.4,K40.8,K40.9,K41,K41.1,K41.2,K41.3,K41.4,K41.8,K41.9,K42,K42.1,K42.2, K42.3,K42.4,K42.8,K42.9,K43,K43.1,K43.2,K43.3,K43.4,K43.8,K43.9,K44,K44.1,K44.2,K44.8,K44.9,K45.1, K45.2,K45.3,K45.4,K45.5,K45.6,K45.8, K45.9,K46,K46.1,K46.2,K46.3,K46.4,K46.5,K46.8,K46.9,K49.1, K49.2,K49.3,K49.4,K49.8,K49.9,K50.1,K50.2,K50.4,K75.1,K75.2, K75.3,K75.4,K75.8,K75.9 |
| **41203** | Diagnosis – main ICD9 | 410,4109,411,4119,412,4129,4140,4148,4149 |
| **41205** | Diagnosis – Secondary ICD9 | 410,4109,411,4119,412,4129,4140,4148,4149 |

**Supplementary Table 2. Summary of 58 evaluated CAD PRS scores in the primary analysis.** Population ranking was based on OR per standard deviation from logistic regression model adjusting for age, sex, PC1-PC4. Individual ranking is based on percentage identification of unexpected early-onset individuals by top 10% of PRS within UK Biobank (n=451,580). PRS scores were further categorized into multi-ancestry, multi-traits and multi-methods. The three categories are not cumulative exhaustive or mutually exclusive. Multi-ancestry scores used multi-ancestry source of variant association or score development population. Multi-traits scores were identified by "incorporation of genetically correlated traits/CAD-related traits/risk factors" or similar wording in their source publications. Multi-methods scores refer to those developed by leveraging multiple scores of the same trait as made clear in source publications.

| PRS | Population Ranking | Individual Ranking | Method | Group | Score development sample size | Citation |
|---|---|---|---|---|---|---|
| **PGS000010** | 53 | 52 | Genome-wide significant variants | Ancestry-Specific | / | Mega JL *et al. Lancet* (2015) |
| **PGS000011** | 55 | 56 | Genome-wide significant variants | Multi-Ancestry | / | Tada H *et al. Eur Heart J* (2015) |
| **PGS000012** | 49 | 49 | LD thinning | Multi-Ancestry | 5883 | Abraham G *et al. Eur Heart J* (2016) |
| **PGS000013** | 33 | 29 | LDpred | Multi-Ancestry | 120280 | Khera AV *et al. Nat Genet* (2018) |

| | | | | | | |
|---|---|---|---|---|---|---|
| **PGS000018** | 29 | 28 | metaGRS | Multi-Ancestry, Multi-Method | 3000 | Inouye M *et al. J Am Coll Cardiol* (2018) |
| **PGS000019** | 47 | 45 | Genome-wide significant variants | Multi-Ancestry | / | Paquette M *et al. J Clin Lipidol* (2017) |
| **PGS000057** | 56 | 54 | Hard thresholding | Multi-Ancestry | / | Natarajan P *et al. Circulation* (2017) |
| **PGS000058** | 37 | 41 | Genome-wide significant variants | Multi-Ancestry | / | Morieri ML *et al. Diabetes Care* (2018) |
| **PGS000059** | 52 | 44 | Literature-derived SNP selection | Multi-Ancestry | / | Hajek C *et al. Circ Genom Precis Med* (2018) |
| **PGS000200** | 57 | 57 | Genome-wide significant variants | Multi-Ancestry | / | Tikkanen E *et al. Arterioscler Thromb Vasc Biol* (2013) |
| **PGS000296** | 35 | 33 | LDpred | Multi-Ancestry | 8766 | Wang M *et al. J Am Coll Cardiol* (2020) |
| **PGS000329** | 3 | 2 | LDpred | Ancestry-Specific | 21813 | Mars N *et al. Nat Med* (2020) |
| **PGS000337** | 6 | 5 | P-value thresholding | Multi-Ancestry | / | Koyama S *et al. Nat Genet* (2020) |

| | | | | | | |
|---|---|---|---|---|---|---|
| **PGS000349** | 58 | 58 | Genome-wide significant variants | Multi-Ancestry | / | Pechlivanis S *et al. BMC Med Genet* (2020) |
| **PGS000746** | 41 | 37 | LD clumping | Multi-Ancestry | 10000 | Gola D *et al. Circ Genom Precis Med* (2020) |
| **PGS000747** | 42 | 32 | LD clumping | Multi-Ancestry | 10000 | Gola D *et al. Circ Genom Precis Med* (2020) |
| **PGS000748** | 46 | 43 | Genome-wide significant variants | Multi-Ancestry | 10000 | Gola D *et al. Circ Genom Precis Med* (2020) |
| **PGS000749** | 44 | 42 | Genome-wide significant variants | Multi-Ancestry | 10000 | Gola D *et al. Circ Genom Precis Med* (2020) |
| **PGS000798** | 40 | 40 | Genome-wide significant variants | Ancestry-Specific | / | Severance LM *et al. J Cardiovasc Comput Tomogr* (2019) |
| **PGS000818** | 45 | 51 | Genome-wide significant variants | Multi-Ancestry | / | Bauer A *et al. Genet Epidemiol* (2021) |
| **PGS000899** | 36 | 38 | SNP selection based on 1) genome-wide significance; 2) linkage disequilibrium | Multi-Ancestry | / | Feitosa MF *et al. Circ Genom Precis Med* (2021) |

| | | | | | | |
|---|---|---|---|---|---|---|
| **PGS000962** | 27 | 34 | snpnet | Ancestry-Specific | 269704 | Tanigawa Y *et al. PLoS Genet* (2022) |
| **PGS001355** | 30 | 35 | AnnoPred | Ancestry-Specific | 92928 | Ye Y *et al. Circ Genom Precis Med* (2021) |
| **PGS001780** | 32 | 30 | PRS-CS | Multi-Ancestry | / | Tamlander M *et al. Commun Biol* (2022) |
| **PGS001839** | 9 | 13 | Penalize regression (bigstatsr) | Ancestry-Specific | 391124 | Privé F *et al. Am J Hum Genet* (2022) |
| **PGS002048** | 8 | 16 | LDpred2 (bigsnpr) | Ancestry-Specific | 391124 | Privé F *et al. Am J Hum Genet* (2022) |
| **PGS002244** | 38 | 36 | LDpred | Multi-Ancestry | 175364 | Mars N *et al. Cell Genom* (2022) |
| **PGS002262** | 51 | 50 | metaPRS | Multi-Ancestry, Multi-Trait | 4855 | Lu X *et al. Eur Heart J* (2022) |
| **PGS002775** | 43 | 39 | Maximum clumping and thresholding (maxCT) | Multi-Ancestry | 38217 | Wong CK *et al. PLoS One* (2022) |
| **PGS002776** | 34 | 31 | Stacked clumping and thresholding (SCT) | Multi-Ancestry | 38217 | Wong CK *et al. PLoS One* (2022) |

| | | | | | | |
|---|---|---|---|---|---|---|
| **PGS002809** | 39 | 47 | Genome-wide significant SNPs | Multi-Ancestry | / | Ahmed R *et al. Int J Cardiol Heart Vasc* (2022) |
| **PGS003355** | 28 | 23 | LDpred | Multi-Ancestry | / | Aragam KG *et al. Nat Genet* (2022) |
| **PGS003356** | 4 | 4 | LDpred | Multi-Ancestry | / | Aragam KG *et al. Nat Genet* (2022) |
| **PGS003438** | 31 | 46 | Genome-wide significant SNPs | Multi-Ancestry | / | Marston NA *et al. JAMA Cardiol* (2023) |
| **PGS003446** | 13 | 7 | PRSice | Multi-Ancestry | / | Tcheandjieu C *et al. Nat Med* (2022) |
| **PGS003725** | 23 | 25 | LDpred2 | Multi-Ancestry | 116649 | Patel AP *et al. Nat Med* (2023) |
| **PGS003726** | 25 | 27 | LDpred2 | Multi-Ancestry | 116649 | Patel AP *et al. Nat Med* (2023) |
| **PGS003727** | 26 | 26 | LDpred2 | Ancestry-Specific | 116649 | Patel AP *et al. Nat Med* (2023) |
| **PGS003866** | 19 | 17 | lassosum2 | Ancestry-Specific | / | Shim I *et al. Nature Communications* (2023) |
| **PGS004196** | 24 | 24 | LASSO | Ancestry-Specific | 200000 | Raben TG *et al. Sci Rep* (2023) |

| | | | | | | |
|---|---|---|---|---|---|---|
| **PGS004197** | 12 | 8 | LASSO | Ancestry-Specific | 200000 | Raben TG *et al. Sci Rep* (2023) |
| **PGS004198** | 21 | 20 | LASSO | Ancestry-Specific | 200000 | Raben TG *et al. Sci Rep* (2023) |
| **PGS004199** | 20 | 21 | LASSO | Ancestry-Specific | 200000 | Raben TG *et al. Sci Rep* (2023) |
| **PGS004200** | 15 | 18 | LASSO | Ancestry-Specific | 200000 | Raben TG *et al. Sci Rep* (2023) |
| **PGS004321** | 50 | 48 | Genome-wide significant SNPs | Multi-Ancestry | / | Marston NA *et al. Circulation* (2019) |
| **PGS004443** | 16 | 11 | LDpred2 | Ancestry-Specific | 174489 | Jung H *et al. Commun Biol* (2024) |
| **PGS004444** | 14 | 6 | LDpred2 | Ancestry-Specific | 174489 | Jung H *et al. Commun Biol* (2024) |
| **PGS004513** | 18 | 15 | RFDiseasemetaPRS | Ancestry-Specific, Multi-Trait | 174489 | Jung H *et al. Commun Biol* (2024) |

| | | | | | | |
|---|---|---|---|---|---|---|
| **PGS004514** | 17 | 12 | RFDiseasemetaPRS | Ancestry-Specific, Multi-Trait | 174489 | Jung H *et al. Commun Biol* (2024) |
| **PGS004595** | 48 | 53 | Genome-wide significant SNPs | Ancestry-Specific | / | Oni-Orisan A *et al. Clin Pharmacol Ther* (2022) |
| **PGS004596** | 54 | 55 | Genome-wide significant SNPs | Multi-Ancestry | / | Peng H *et al. Nutrients* (2023) |
| **PGS004696** | 2 | 3 | PRS-CSx | Multi-Ancestry | 87724 | Smith JL *et al. Circ Genom Precis Med* (2024) |
| **PGS004697** | 1 | 1 | PRS-CSx | Multi-Ancestry | 56359 | Smith JL *et al. Circ Genom Precis Med* (2024) |
| **PGS004698** | 7 | 9 | Pruning and Thresholding | Multi-Ancestry | 87724 | Smith JL *et al. Circ Genom Precis Med* (2024) |
| **PGS004743** | 10 | 14 | PRSmix | Ancestry-Specific, Multi-Method | 29863 | Truong B et al., *Cell Genom* (2024) |
| **PGS004744** | 22 | 22 | PRSmix | Ancestry-Specific, Multi-Method | 35350 | Truong B et al., *Cell Genom* (2024) |

| | | | | | | |
|---|---|---|---|---|---|---|
| **PGS004745** | 5 | 19 | PRSmix | Ancestry-Specific, Multi-Method | 29863 | Truong B et al., *Cell Genom* (2024) |
| **PGS004746** | 11 | 10 | PRSmixPlus | Ancestry-Specific, Multi-Method | 35350 | Truong B et al., *Cell Genom* (2024) |

Information on scores (method, score development sample size and citation) extracted from PGS catalog (https://www.pgscatalog.org).

**Supplementary Table 3. Baseline characteristics of UK Biobank participants defined as benchmark high genetic risk CAD population in sensitivity analysis 1-4.** Definitions of benchmark cohorts differ in sensitivity analysis 1-4 (**Table 4).** In sensitivity analysis 1, benchmark cohort was defined in the same way as the primary analysis, except for differential early onset cutoff age (60 years old) for females. In sensitivity analysis 2, benchmark cohort was defined as those in the primary analysis benchmark cohort also having family history of heart disease in both parents. In sensitivity analysis 3, benchmark cohort was defined as CAD patients with family history of heart disease in both parents. In sensitivity analysis 4, benchmark cohort is defined by lack of standard modifiable risk factors and onset age < 55 years old.

| Metric [a] | Sensitivity Analysis 1 (n=1699) | Sensitivity Analysis 2 (n=109) | Sensitivity Analysis 3 (n=6534) | Sensitivity Analysis 4 (n=173) |
|---|---|---|---|---|
| **Male** | 612 (36.0) | 42 (38.5) | 3791 (59.8) | 84 (48.6) |
| **Age (years)** | 47.90 (5.16) | 47.07 (4.37) | 60.65 (6.26) | 47.28 (5.06) |
| **British White Ancestry** | 1361 (80.1) | 83 (76.1) | 5430 (85.7) | 135 (78.0) |
| **Townsend Deprivation Index [b]** | -0.43 (3.37) | 0.53 (3.60) | -1.01 (3.23) | -0.47 (3.41) |
| **Pooled Cohort Equation Risk** | 0.02 (0.01) | 0.02 (0.01) | 0.13 (0.08) | 0.02 (0.02) |
| **LE8 Diet Score [c]** | 50.16 (32.25) | 52.90 (33.88) | 51.50 (32.58) | 54.69 (33.16) |
| **LE8 PA Score** | 31.15 (28.55) | 31.68 (29.64) | 30.40 (28.37) | 31.60 (26.70) |
| **LE8 Smoking Score** | 85.43 (30.79) | 85.28 (32.75) | 80.78 (31.82) | 94.14 (16.14) |
| **LE8 Sleep Health Points** | 86.10 (22.38) | 85.98 (23.51) | 86.51 (22.31) | 89.51 (18.81) |

| | | | | |
|---|---|---|---|---|
| **LE8 BMI Points** | 63.90 (30.92) | 61.78 (31.55) | 58.11 (29.90) | 71.05 (28.58) |
| **LE8 Blood Lipid Points** | 49.33 (28.38) | 53.27 (28.01) | 51.32 (30.42) | 75.43 (22.87) |
| **LE8 HbA1c Score** | 94.92 (14.59) | 91.40 (19.06) | 79.44 (28.97) | 98.77 (6.94) |
| **LE8 BP Score** | 53.34 (26.63) | 52.94 (28.05) | 40.72 (26.18) | 68.36 (21.55) |
| **LE8 Composite Score** | 64.29 (10.57) | 64.40 (11.79) | 59.85 (11.71) | 72.94 (9.53) |
| **C Reactive Protein** | 3.16 (5.04) | 3.67 (5.34) | 3.27 (5.24) | 2.78 (4.88) |
| **Family Heart Disease History** | | | | |
| ***Both Parents*** | 179 (10.5) | 109 (100.0) | 6534 (100.0) | 16 (9.2) |
| ***Father*** | 425 (25.0) | 0 (0.00) | 0 (0.00) | 37 (21.4) |
| ***Mother*** | 218 (12.8) | 0 (0.00) | 0 (0.00) | 18 (10.4) |
| ***No History*** | 877 (51.6) | 0 (0.00) | 0 (0.00) | 102 (59.0) |
| **Familial Hypercholesterolemia Variant Carrier** | 6 (0.35) | 0 (0.00) | 73 (1.12) | 0 (0.00) |
| **Thrombophilia Variant [d]** | | | | |
| ***High Risk*** | 40 (2.35) | 6 (5.50) | 184 (0.55) | 9 (5.2) |
| ***Low Risk*** | 26 (1.53) | 2 (1.83) | 69 (0.21) | 1 (0.58) |

[a] Metrics are presented as mean (standard deviation) for continuous variables and n (%) for categorical variables

[b] Higher Townsend Deprivation Index indicates greater degree of deprivation.

[c] Higher Life's Essential 8 score indicates more favourable profile.

[d] High-risk variant = High-risk non-synonymous Human Gene Mutation Database variants in the 3 anticoagulant genes SERPINC1, PROC, and PROS1. Low-risk variant = Low-risk non-synonymous non-Human Gene Mutation Database variants in the 3 anticoagulant genes SERPINC1, PROC, and PROS1.

**Supplementary Table 4. Summary of 58 evaluated CAD PRS scores in sensitivity analysis 1-3.** Population ranking for primary analysis and sensitivity analysis 1-3 was based on OR per standard deviation from logistic regression model adjusting for age, sex, PC1-PC4. Individual ranking is based on percentage identification of benchmark cohort by top 10% of PRS within UK Biobank (n=451,580). Definitions of benchmark cohorts differ in sensitivity analysis 1-3 (**Table 4**). In sensitivity analysis 1, benchmark cohort was defined in the same way as the primary analysis, except for differential early onset cutoff age (60 years old) for females. In sensitivity analysis 2, benchmark cohort was defined as those in the primary analysis benchmark cohort also having family history of heart disease in both parents. In sensitivity analysis 3, benchmark cohort was defined as CAD patients with family history of heart disease in both parents. Individual rankings of sensitivity analysis 1-3 correlated well with that of the primary analysis by Spearman's rho of 0.99, 0.90 and 0.94 respectively (**Table 4**).

| PRS | Population Ranking | Individual Ranking Primary Analysis | Individual Ranking Sensitivity Analysis 1 | Individual Ranking Sensitivity Analysis 2 | Individual Ranking Sensitivity Analysis 3 |
|---|---|---|---|---|---|
| **PGS000010** | 51 | 53 | 53 | 48 | 52 |
| **PGS000011** | 55 | 37 | 55 | 43 | 53 |
| **PGS000012** | 53 | 54 | 48 | 40 | 50 |
| **PGS000013** | 31 | 30 | 30 | 31 | 33 |
| **PGS000018** | 32 | 33 | 28 | 37 | 32 |
| **PGS000019** | 42 | 38 | 47 | 49 | 47 |
| **PGS000057** | 57 | 48 | 56 | 57 | 55 |

| | | | | | |
|---|---|---|---|---|---|
| **PGS000058** | 36 | 39 | 37 | 51 | 30 |
| **PGS000059** | 50 | 27 | 43 | 41 | 51 |
| **PGS000200** | 56 | 57 | 57 | 52 | 56 |
| **PGS000296** | 37 | 31 | 34 | 32 | 36 |
| **PGS000329** | 2 | 2 | 3 | 2 | 2 |
| **PGS000337** | 11 | 6 | 5 | 10 | 8 |
| **PGS000349** | 58 | 58 | 58 | 42 | 58 |
| **PGS000746** | 41 | 40 | 42 | 50 | 41 |
| **PGS000747** | 47 | 25 | 36 | 25 | 44 |
| **PGS000748** | 48 | 41 | 40 | 35 | 46 |
| **PGS000749** | 46 | 45 | 44 | 26 | 45 |
| **PGS000798** | 40 | 28 | 45 | 46 | 40 |
| **PGS000818** | 44 | 34 | 50 | 53 | 43 |
| **PGS000899** | 35 | 26 | 39 | 55 | 35 |
| **PGS000962** | 27 | 22 | 31 | 19 | 27 |
| **PGS001355** | 29 | 49 | 33 | 33 | 31 |
| **PGS001780** | 30 | 23 | 29 | 23 | 34 |
| **PGS001839** | 6 | 13 | 9 | 12 | 7 |

| | | | | | |
|---|---|---|---|---|---|
| **PGS002048** | 5 | 15 | 12 | 14 | 6 |
| **PGS002244** | 38 | 42 | 35 | 38 | 38 |
| **PGS002262** | 52 | 55 | 52 | 58 | 57 |
| **PGS002775** | 43 | 46 | 38 | 54 | 42 |
| **PGS002776** | 34 | 35 | 32 | 34 | 37 |
| **PGS002809** | 39 | 43 | 46 | 44 | 39 |
| **PGS003355** | 28 | 21 | 25 | 36 | 29 |
| **PGS003356** | 7 | 7 | 6 | 5 | 5 |
| **PGS003438** | 33 | 36 | 41 | 39 | 28 |
| **PGS003446** | 18 | 9 | 7 | 6 | 14 |
| **PGS003725** | 25 | 50 | 23 | 21 | 22 |
| **PGS003726** | 24 | 51 | 24 | 27 | 24 |
| **PGS003727** | 26 | 47 | 26 | 24 | 26 |
| **PGS003866** | 21 | 3 | 19 | 17 | 20 |
| **PGS004196** | 23 | 18 | 27 | 28 | 21 |
| **PGS004197** | 8 | 8 | 10 | 8 | 4 |
| **PGS004198** | 20 | 16 | 20 | 29 | 16 |
| **PGS004199** | 19 | 14 | 21 | 18 | 17 |

| | | | | | |
|---|---|---|---|---|---|
| **PGS004200** | 14 | 17 | 18 | 16 | 11 |
| **PGS004321** | 49 | 44 | 49 | 47 | 49 |
| **PGS004443** | 9 | 10 | 14 | 30 | 15 |
| **PGS004444** | 3 | 1 | 4 | 9 | 18 |
| **PGS004513** | 17 | 20 | 17 | 20 | 19 |
| **PGS004514** | 13 | 11 | 15 | 7 | 23 |
| **PGS004595** | 45 | 56 | 51 | 56 | 48 |
| **PGS004596** | 54 | 52 | 54 | 45 | 54 |
| **PGS004696** | 4 | 4 | 2 | 3 | 3 |
| **PGS004697** | 1 | 5 | 1 | 1 | 1 |
| **PGS004698** | 15 | 12 | 8 | 15 | 13 |
| **PGS004743** | 12 | 19 | 13 | 13 | 9 |
| **PGS004744** | 22 | 32 | 22 | 22 | 25 |
| **PGS004745** | 10 | 24 | 16 | 11 | 10 |
| **PGS004746** | 16 | 29 | 11 | 4 | 12 |

**Supplementary Table 5. Individuals with early onset unexpected CAD stratified by how often they are captured by 58 CAD PRSs in sensitivity analysis 1.** The "Consistently Low-PRS" group is defined as individuals with early onset unexpected CAD who have never been identified as top 10% risk by any of the 58 PRS scores. The "Consistently High-PRS" group is defined as individuals with early onset unexpected CAD who have been identified as top 10% risk by more than 20 out of the 58 PRSs.

| **Metric [a]** | **Consistently Low-PRS (n=207)** | **Consistently High-PRS (n=319)** | **P-value [b]** |
|---|---|---|---|
| **Male** | 70 (33.8) | 139 (43.6) | 0.032 |
| **Age (years)** | 48.42 (5.41) | 47.53 (5.04) | 0.060 |
| **Townsend Deprivation Index [c]** | 0.02 (3.37) | -0.70 (3.06) | 0.014 |
| **LE8 Diet Score [d]** | 51.68 (34.43) | 52.15 (30.36) | 0.87 |
| **LE8 PA Score** | 32.34 (30.16) | 32.41 (28.51) | 0.98 |
| **LE8 Smoking Score** | 79.31 (35.99) | 85.13 (30.81) | 0.062 |
| **LE8 Sleep Health Points** | 86.29 (21.24) | 86.75 (22.29) | 0.82 |
| **LE8 BMI Points** | 64.34 (31.85) | 64.37 (30.65) | 0.99 |
| **LE8 Blood Lipid Points** | 54.92 (29.39) | 41.03 (25.88) | < 0.001 |
| **LE8 HbA1c Score** | 95.74 (13.33) | 92.93 (16.50) | 0.036 |
| **LE8 BP Score** | 56.85 (28.53) | 51.21 (27.04) | 0.027 |
| **LE8 Composite Score** | 65.18 (11.08) | 63.25 (10.21) | 0.049 |

| | | | |
|---|---|---|---|
| **Race** | | | 0.13 |
| ***White*** | 188 (90.8) | 304 (95.3) | |
| ***Black*** | 2 (1.0) | 3 (0.9) | |
| ***South Asian*** | 9 (4.3) | 8 (2.5) | |
| ***Other*** | 8 (3.9) | 4 (1.3) | |
| **Alcohol intake frequency** | | | 0.11 |
| ***1-2x/wk*** | 57 (27.5) | 95 (29.8) | |
| ***1-3x/mo*** | 19 (9.2) | 40 (12.5) | |
| ***3-4x/wk*** | 36 (17.4) | 63 (19.7) | |
| ***Daily or almost Daily*** | 27 (13.0) | 51 (16.0) | |
| ***Never*** | 37 (17.9) | 33 (10.3) | |
| ***Special Occasions Only*** | 31 (15.0) | 37 (11.6) | |
| **Smoking** | | | 0.26 |
| ***Never*** | 115 (55.6) | 185 (58.0) | |
| ***Previous*** | 63 (30.4) | 104 (32.6) | |
| ***Current*** | 29 (14.0) | 30 (9.4) | |
| **Chronic Kidney Disease** | 8 (3.9) | 8 (2.5) | 0.53 |
| **Rheumatoid arthritis** | 1 (0.5) | 11 (3.4) | 0.033 |

| | | | |
|---|---|---|---|
| **Pooled Cohort Equation Risk** | 0.02 (0.01) | 0.03 (0.01) | 0.0015 |
| **Lipoprotein A** | 34.93 (39.08) | 73.06 (59.32) | < 0.001 |
| **C reactive protein** | 2.65 (3.92) | 3.14 (5.46) | 0.22 |
| **Age of CAD Onset** | 52.76 (5.44) | 52.01 (4.98) | 0.11 |
| **Family Heart Disease History** | | | < 0.001 |
| ***Both Parents*** | 17 (8.2) | 47 (14.7) | |
| ***Father*** | 37 (17.9) | 100 (31.3) | |
| ***Mother*** | 26 (12.6) | 48 (15.0) | |
| ***No History*** | 127 (61.4) | 124 (38.9) | |
| **Familial Hypercholesterolemia Variant Carrier** | 1 (0.48) | 1 (0.31) | 1.00 |
| **Thrombophilia Variant** [e] | | | 0.36 |
| ***High Risk*** | 5 (2.42) | 6 (1.88) | |
| ***Low Risk*** | 2 (0.97) | 8 (2.51) | |

[a] Metrics are represented as mean (standard deviation) for continuous variables and % (n) for categorical variables.

[b] P values calculated with 2-sample t-test for continuous traits and Chi-squared or Fisher exact test for categorical traits.

[c] Higher Townsend Deprivation Index indicates greater degree of deprivation.

[d] Higher Life's Essential 8 score indicates more favourable profile.

[e] High-risk variant = High-risk non-synonymous Human Gene Mutation Database variants in the 3 anticoagulant genes SERPINC1, PROC, and PROS1. Low-risk variant = Low-risk non-synonymous non-Human Gene Mutation Database variants in the 3 anticoagulant genes SERPINC1, PROC, and PROS1.

**Supplementary Table 6. Individuals with early onset unexpected CAD stratified by how often they are captured by 58 CAD PRSs in sensitivity analysis 3.** The “Consistently Low-PRS” group is defined as individuals with early onset unexpected CAD who have never been identified as top 10% risk by any of the 58 PRS scores. The “Consistently High-PRS” group is defined as individuals with early onset unexpected CAD who have been identified as top 10% risk by more than 20 out of the 58 PRSs. Family history of heart disease is not compared for this group since by definition all benchmark cohort has family history of heart disease in both parents.

| Metric [a] | Consistently Low-PRS (n=327) | Consistently High-PRS (n=831) | P-value [b] |
|---|---|---|---|
| **Male** | 186 (56.9) | 549 (66.1) | 0.0043 |
| **Age (years)** | 61.14 (6.18) | 59.83 (6.40) | 0.0013 |
| **Townsend Deprivation Index [c]** | -1.06 (3.19) | -1.12 (3.27) | 0.78 |
| **LE8 Diet Score [d]** | 52.42 (32.50) | 51.64 (32.21) | 0.72 |
| **LE8 PA Score** | 31.81 (29.20) | 31.39 (28.99) | 0.83 |
| **LE8 Smoking Score** | 80.40 (31.24) | 77.04 (35.02) | 0.12 |
| **LE8 Sleep Health Points** | 85.03 (23.01) | 87.16 (21.59) | 0.16 |
| **LE8 BMI Points** | 58.39 (30.41) | 57.78 (29.21) | 0.76 |
| **LE8 Blood Lipid Points** | 50.71 (28.85) | 51.53 (29.52) | 0.67 |
| **LE8 HbA1c Score** | 81.29 (27.43) | 79.71 (28.37) | 0.39 |
| **LE8 BP Score** | 39.37 (25.23) | 39.65 (26.18) | 0.87 |

| | | | |
|---|---|---|---|
| **LE8 Composite Score** | 59.93 (11.74) | 59.49 (11.84) | 0.58 |
| **Race** | | | 0.0023 |
| ***White*** | 299 (91.4) | 800 (96.3) | |
| ***Black*** | 2 (0.6) | 0 (0.0) | |
| ***South Asian*** | 21 (6.4) | 24 (2.9) | |
| ***Other*** | 5 (1.5) | 7 (0.8) | |
| **Alcohol intake frequency** | | | 0.0012 |
| ***1-2x/wk*** | 84 (25.7) | 213 (25.6) | |
| ***1-3x/mo*** | 30 (9.2) | 109 (13.1) | |
| ***3-4x/wk*** | 48 (14.7) | 189 (22.7) | |
| ***Daily or almost Daily*** | 78 (23.9) | 143 (17.2) | |
| ***Never*** | 43 (13.1) | 75 (9.0) | |
| ***Special Occasions Only*** | 44 (13.5) | 102 (12.3) | |
| **Smoking** | | | 0.059 |
| ***Never*** | 78 (59.1) | 135 (59.2) | |
| ***Previous*** | 33 (25.0) | 73 (32.0) | |
| ***Current*** | 21 (15.9) | 20 (8.8) | |
| **Chronic Kidney Disease** | 38 (11.6) | 87 (10.5) | 0.64 |

| | | | |
|---|---|---|---|
| **Rheumatoid arthritis** | 9 (2.8) | 41 (4.9) | 0.14 |
| **Pooled Cohort Equation Risk** | 0.13 (0.09) | 0.13 (0.08) | 0.79 |
| **Lipoprotein A** | 39.79 (45.13) | 61.77 (57.99) | < 0.001 |
| **C reactive protein** | 3.39 (5.07) | 3.02 (4.77) | 0.25 |
| **Age of CAD Onset** | 66.04 (8.32) | 59.90 (8.46) | < 0.001 |
| **Familial Hypercholesterolemia Variant Carrier** | 1 (0.31) | 6 (0.12) | 0.68 |
| **Thrombophilia Variant** [e] | | | 0.44 |
| ***High Risk*** | 8 (2.45) | 28 (3.37) | |
| ***Low Risk*** | 5 (1.53) | 8 (0.96) | |

[a] Metrics are represented as mean (standard deviation) for continuous variables and % (n) for categorical variables.

[b] P values calculated with 2-sample t-test for continuous traits and Chi-squared or Fisher exact test for categorical traits.

[c] Higher Townsend Deprivation Index indicates greater degree of deprivation.

[d] Higher Life's Essential 8 score indicates more favourable profile.

[e] High-risk variant = High-risk non-synonymous Human Gene Mutation Database variants in the 3 anticoagulant genes SERPINC1, PROC, and PROS1. Low-risk variant = Low-risk non-synonymous non-Human Gene Mutation Database variants in the 3 anticoagulant genes SERPINC1, PROC, and PROS1.

**Supplementary Table 7. Definition of unexpected CAD in sensitivity analysis 4 and 5.** The unexpected CAD cohort was identified as patients with none of the following risk factors defined by the listed criteria at baseline in UKBB (sensitivity analysis 4); or patients with none of the following risk factors defined by the listed criteria, except for Diabetes Mellitus medication status, at diagnosis of CAD in MGBB linked electronic health records (sensitivity analysis 5). Both cohorts were further restricted to individuals with CAD event diagnosed at least 6 months before any risk factor.

| **Risk Factor** | **Definition** |
|---|---|
| **Hypertension** | SBP ≥ 140 mm Hg ***or***<br>DBP ≥ 90 mm Hg ***or***<br>On Hypertension Medication |
| **Diabetes Mellitus** | Hba1c ≥ 6.5% ***or***<br>On Diabetes Mellitus Medication [a] |
| **Hypercholesterolemia** | TC > 5.5 mmol/L ***or***<br>LDL-C > 3.5 mmol/L ***or***<br>On Hypercholesterolemia Medication |
| **Cigarette Smoking** | Current Smoker |

Adapted from Figtree Gemma et al[1]. DBP = diastolic blood pressure; HbA1c = glycosylated hemoglobin; LDL-C = low-density lipoprotein cholesterol; SBP = systolic blood pressure; TC = total cholesterol.

[a] Diabetes Mellitus medication status was not used in benchmark cohort definition of sensitivity analysis 5.

**Supplementary Table 8. Summary of 58 evaluated CAD PRS scores in sensitivity analysis 4.** Population ranking was based on HR per standard deviation from Cox proportional hazard model accounting for only person-time without the 4 defining risk factors of unexpected CAD adjusting for age, sex, PC1-PC4. Individual ranking is based on percentage identification of unexpected early-onset individuals by top 10% of PRS within UK Biobank (n=451,580). PRS scores were further categorized into multi-ancestry, multi-traits and multi-methods. The three categories are not cumulative exhaustive or mutually exclusive. Multi-ancestry scores used multi-ancestry source of variant association or score development population. Multi-traits scores were identified by "incorporation of genetically correlated traits/CAD-related traits/risk factors" or similar wording in their source publications. Multi-methods scores refer to those developed by leveraging multiple scores of the same trait as made clear in source publications.

| PRS | Population Ranking | Individual Ranking | Method | Group | Score development sample size | Citation |
|---|---|---|---|---|---|---|
| **PGS000010** | 51 | 53 | Genome-wide significant variants | Ancestry-Specific | / | Mega JL *et al. Lancet* (2015) |
| **PGS000011** | 55 | 37 | Genome-wide significant variants | Multi-Ancestry | / | Tada H *et al. Eur Heart J* (2015) |
| **PGS000012** | 53 | 54 | LD thinning | Multi-Ancestry | 5883 | Abraham G *et al. Eur Heart J* (2016) |

| | | | | | | |
|---|---|---|---|---|---|---|
| **PGS000013** | 31 | 30 | LDpred | Multi-Ancestry | 120280 | Khera AV *et al. Nat Genet* (2018) |
| **PGS000018** | 32 | 33 | metaGRS | Multi-Ancestry, Multi-Method | 3000 | Inouye M *et al. J Am Coll Cardiol* (2018) |
| **PGS000019** | 42 | 38 | Genome-wide significant variants | Multi-Ancestry | / | Paquette M *et al. J Clin Lipidol* (2017) |
| **PGS000057** | 57 | 48 | Hard thresholding | Multi-Ancestry | / | Natarajan P *et al. Circulation* (2017) |
| **PGS000058** | 36 | 39 | Genome-wide significant variants | Multi-Ancestry | / | Morieri ML *et al. Diabetes Care* (2018) |
| **PGS000059** | 50 | 27 | Literature-derived SNP selection | Multi-Ancestry | / | Hajek C *et al. Circ Genom Precis Med* (2018) |
| **PGS000200** | 56 | 57 | Genome-wide significant variants | Multi-Ancestry | / | Tikkanen E *et al. Arterioscler Thromb Vasc Biol* (2013) |
| **PGS000296** | 37 | 31 | LDpred | Multi-Ancestry | 8766 | Wang M *et al. J Am Coll Cardiol* (2020) |
| **PGS000329** | 2 | 2 | LDpred | Ancestry-Specific | 21813 | Mars N *et al. Nat Med* (2020) |

| | | | | | | |
|---|---|---|---|---|---|---|
| **PGS000337** | 11 | 6 | P-value thresholding | Multi-Ancestry | / | Koyama S *et al. Nat Genet* (2020) |
| **PGS000349** | 58 | 58 | Genome-wide significant variants | Multi-Ancestry | / | Pechlivanis S *et al. BMC Med Genet* (2020) |
| **PGS000746** | 41 | 40 | LD clumping | Multi-Ancestry | 10000 | Gola D *et al. Circ Genom Precis Med* (2020) |
| **PGS000747** | 47 | 25 | LD clumping | Multi-Ancestry | 10000 | Gola D *et al. Circ Genom Precis Med* (2020) |
| **PGS000748** | 48 | 41 | Genome-wide significant variants | Multi-Ancestry | 10000 | Gola D *et al. Circ Genom Precis Med* (2020) |
| **PGS000749** | 46 | 45 | Genome-wide significant variants | Multi-Ancestry | 10000 | Gola D *et al. Circ Genom Precis Med* (2020) |
| **PGS000798** | 40 | 28 | Genome-wide significant variants | Ancestry-Specific | / | Severance LM *et al. J Cardiovasc Comput Tomogr* (2019) |
| **PGS000818** | 44 | 34 | Genome-wide significant variants | Multi-Ancestry | / | Bauer A *et al. Genet Epidemiol* (2021) |

| | | | | | | |
|---|---|---|---|---|---|---|
| **PGS000899** | 35 | 26 | SNP selection based on 1) genome-wide significance; 2) linkage disequilibrium | Multi-Ancestry | / | Feitosa MF *et al. Circ Genom Precis Med* (2021) |
| **PGS000962** | 27 | 22 | snpnet | Ancestry-Specific | 269704 | Tanigawa Y *et al. PLoS Genet* (2022) |
| **PGS001355** | 29 | 49 | AnnoPred | Ancestry-Specific | 92928 | Ye Y *et al. Circ Genom Precis Med* (2021) |
| **PGS001780** | 30 | 23 | PRS-CS | Multi-Ancestry | / | Tamlander M *et al. Commun Biol* (2022) |
| **PGS001839** | 6 | 13 | Penalize regression (bigstatsr) | Ancestry-Specific | 391124 | Privé F *et al. Am J Hum Genet* (2022) |
| **PGS002048** | 5 | 15 | LDpred2 (bigsnpr) | Ancestry-Specific | 391124 | Privé F *et al. Am J Hum Genet* (2022) |
| **PGS002244** | 38 | 42 | LDpred | Multi-Ancestry | 175364 | Mars N *et al. Cell Genom* (2022) |
| **PGS002262** | 52 | 55 | metaPRS | Multi-Ancestry, Multi-Trait | 4855 | Lu X *et al. Eur Heart J* (2022) |
| **PGS002775** | 43 | 46 | Maximum clumping and thresholding (maxCT) | Multi-Ancestry | 38217 | Wong CK *et al. PLoS One* (2022) |

| | | | | | | |
|---|---|---|---|---|---|---|
| **PGS002776** | 34 | 35 | Stacked clumping and thresholding (SCT) | Multi-Ancestry | 38217 | Wong CK *et al. PLoS One* (2022) |
| **PGS002809** | 39 | 43 | Genome-wide significant SNPs | Multi-Ancestry | / | Ahmed R *et al. Int J Cardiol Heart Vasc* (2022) |
| **PGS003355** | 28 | 21 | LDpred | Multi-Ancestry | / | Aragam KG *et al. Nat Genet* (2022) |
| **PGS003356** | 7 | 7 | LDpred | Multi-Ancestry | / | Aragam KG *et al. Nat Genet* (2022) |
| **PGS003438** | 33 | 36 | Genome-wide significant SNPs | Multi-Ancestry | / | Marston NA *et al. JAMA Cardiol* (2023) |
| **PGS003446** | 18 | 9 | PRSice | Multi-Ancestry | / | Tcheandjieu C *et al. Nat Med* (2022) |
| **PGS003725** | 25 | 50 | LDpred2 | Multi-Ancestry | 116649 | Patel AP *et al. Nat Med* (2023) |
| **PGS003726** | 24 | 51 | LDpred2 | Multi-Ancestry | 116649 | Patel AP *et al. Nat Med* (2023) |
| **PGS003727** | 26 | 47 | LDpred2 | Ancestry-Specific | 116649 | Patel AP *et al. Nat Med* (2023) |
| **PGS003866** | 21 | 3 | lassosum2 | Ancestry-Specific | / | Shim I *et al. Nature Communications* (2023) |

| | | | | | | |
|---|---|---|---|---|---|---|
| **PGS004196** | 23 | 18 | LASSO | Ancestry-Specific | 200000 | Raben TG *et al. Sci Rep* (2023) |
| **PGS004197** | 8 | 8 | LASSO | Ancestry-Specific | 200000 | Raben TG *et al. Sci Rep* (2023) |
| **PGS004198** | 20 | 16 | LASSO | Ancestry-Specific | 200000 | Raben TG *et al. Sci Rep* (2023) |
| **PGS004199** | 19 | 14 | LASSO | Ancestry-Specific | 200000 | Raben TG *et al. Sci Rep* (2023) |
| **PGS004200** | 14 | 17 | LASSO | Ancestry-Specific | 200000 | Raben TG *et al. Sci Rep* (2023) |
| **PGS004321** | 49 | 44 | Genome-wide significant SNPs | Multi-Ancestry | / | Marston NA *et al. Circulation* (2019) |
| **PGS004443** | 9 | 10 | LDpred2 | Ancestry-Specific | 174489 | Jung H *et al. Commun Biol* (2024) |
| **PGS004444** | 3 | 1 | LDpred2 | Ancestry-Specific | 174489 | Jung H *et al. Commun Biol* (2024) |
| **PGS004513** | 17 | 20 | RFDiseasemetaPRS | Ancestry-Specific, | 174489 | Jung H *et al. Commun Biol* (2024) |

| | | | | | | |
|---|---|---|---|---|---|---|
| | | | | Multi-Trait | | |
| **PGS004514** | 13 | 11 | RFDiseasemetaPRS | Ancestry-Specific, Multi-Trait | 174489 | Jung H *et al. Commun Biol* (2024) |
| **PGS004595** | 45 | 56 | Genome-wide significant SNPs | Ancestry-Specific | / | Oni-Orisan A *et al. Clin Pharmacol Ther* (2022) |
| **PGS004596** | 54 | 52 | Genome-wide significant SNPs | Multi-Ancestry | / | Peng H *et al. Nutrients* (2023) |
| **PGS004696** | 4 | 4 | PRS-CSx | Multi-Ancestry | 87724 | Smith JL *et al. Circ Genom Precis Med* (2024) |
| **PGS004697** | 1 | 5 | PRS-CSx | Multi-Ancestry | 56359 | Smith JL *et al. Circ Genom Precis Med* (2024) |
| **PGS004698** | 15 | 12 | Pruning and Thresholding | Multi-Ancestry | 87724 | Smith JL *et al. Circ Genom Precis Med* (2024) |
| **PGS004743** | 12 | 19 | PRSmix | Ancestry-Specific, Multi-Method | 29863 | Truong B et al., *Cell Genom* (2024) |
| **PGS004744** | 22 | 32 | PRSmix | Ancestry-Specific, | 35350 | Truong B et al., *Cell Genom* (2024) |

| | | | | | | |
|---|---|---|---|---|---|---|
| | | | | Multi-Method | | |
| **PGS004745** | 10 | 24 | PRSmix | Ancestry-Specific, Multi-Method | 29863 | Truong B et al., *Cell Genom* (2024) |
| **PGS004746** | 16 | 29 | PRSmixPlus | Ancestry-Specific, Multi-Method | 35350 | Truong B et al., *Cell Genom* (2024) |

Information on scores (method, score development sample size and citation) extracted from PGS catalog (https://www.pgscatalog.org).

**Supplementary Table 9. Individuals with early onset unexpected CAD stratified by how often they are captured by 58 CAD PRSs in sensitivity analysis 4.** The "Consistently Low-PRS" group is defined as individuals with early onset unexpected CAD who have never been identified as top 10% risk by any of the 58 PRS scores. The "Consistently High-PRS" group is defined as individuals with early onset unexpected CAD who have been identified as top 10% risk by more than 20 out of the 58 PRSs. Smoking status is not compared since by definition all benchmark cohort are not current smokers. Familial Hypercholesterolemia variant status is not compared since by definition all benchmark cohort has no familial hypercholesterolemia.

| **Metric [a]** | **Consistently Low-PRS (n=26)** | **Consistently High-PRS (n=22)** | **P-value [b]** |
|---|---|---|---|
| **Male** | 16 (61.5) | 9 (40.9) | 0.26 |
| **Age (years)** | 46.88 (4.13) | 47.09 (4.00) | 0.86 |
| **Townsend Deprivation Index [c]** | -0.61 (3.19) | -1.02 (3.47) | 0.67 |
| **LE8 Diet Score [d]** | 56.30 (37.88) | 52.50 (29.59) | 0.71 |
| **LE8 PA Score** | 26.09 (17.51) | 34.55 (27.73) | 0.23 |
| **LE8 Smoking Score** | 93.48 (18.80) | 92.05 (19.50) | 0.80 |
| **LE8 Sleep Health Points** | 93.48 (12.65) | 93.18 (15.85) | 0.95 |
| **LE8 BMI Points** | 62.61 (33.02) | 73.18 (33.68) | 0.29 |
| **LE8 Blood Lipid Points** | 80.87 (22.95) | 60.00 (26.19) | 0.0070 |
| **LE8 HbA1c Score** | 98.26 (8.34) | 100.00 (0.00) | 0.33 |

| | | | |
|---|---|---|---|
| **LE8 BP Score** | 69.57 (22.56) | 70.45 (21.32) | 0.89 |
| **LE8 Composite Score** | 72.58 (11.08) | 71.99 (9.10) | 0.85 |
| **Race** | | | 0.055 |
| ***White*** | 22 (84.6) | 20 (90.9) | |
| **Alcohol intake frequency** | | | 0.32 |
| ***1-2x/wk*** | 8 (32.0) | 8 (36.4) | |
| ***1-3x/mo*** | 2 (8.0) | 4 (18.2) | |
| ***3-4x/wk*** | 4 (16.0) | 7 (31.8) | |
| ***Daily or almost Daily*** | 2 (8.0) | 0 (0.0) | |
| ***Never*** | 4 (16.0) | 1 (4.5) | |
| ***Special Occasions Only*** | 5 (20.0) | 2 (9.1) | |
| **Chronic Kidney Disease** | 1 (3.8) | 0 (0.0) | 1.00 |
| **Rheumatoid arthritis** | 0 (0.0) | 0 (0.0) | 0.56 |
| **Pooled Cohort Equation Risk** | 0.02 (0.01) | 0.02 (0.02) | 0.65 |
| **Lipoprotein A** | 32.83 (37.53) | 88.98 (61.87) | 0.0015 |
| **C reactive protein** | 2.79 (5.04) | 2.75 (5.85) | 0.98 |
| **Age of CAD Onset** | 49.16 (6.13) | 49.44 (3.84) | 0.84 |
| **Family Heart Disease History** | | | 0.044 |

| | | | |
|---|---|---|---|
| ***Both Parents*** | 2 (7.7) | 3 (13.6) | |
| ***Father*** | 3 (11.5) | 9 (40.9) | |
| ***Mother*** | 2 (7.7) | 2 (9.1) | |
| ***No History*** | 19 (73.1) | 8 (36.4) | |
| **Thrombophilia Variant [e]** | | | 1.00 |
| ***High Risk*** | 2 (7.69) | 0 (0.0) | |
| ***Low Risk*** | 0 (0.0) | 0 (0.0) | |

[a] Metrics are represented as mean (standard deviation) for continuous variables and % (n) for categorical variables.

[b] P values calculated with 2-sample t-test for continuous traits and Chi-squared or Fisher exact test for categorical traits.

[c] Higher Townsend Deprivation Index indicates greater degree of deprivation.

[d] Higher Life’s Essential 8 score indicates more favourable profile.

[e] High-risk variant = High-risk non-synonymous Human Gene Mutation Database variants in the 3 anticoagulant genes SERPINC1, PROC, and PROS1. Low-risk variant = Low-risk non-synonymous non-Human Gene Mutation Database variants in the 3 anticoagulant genes SERPINC1, PROC, and PROS1.

**Supplementary Table 10. Summary of 58 evaluated CAD PRS scores in sensitivity analysis 5 using MGBB data.** Population ranking was based on OR per standard deviation from logistic regression model adjusting for age, sex and categorical ancestry. Individual ranking is based on percentage identification of unexpected early-onset individuals by top 10% of PRS within MGBB (n=49,744). PRS scores were further categorized into multi-ancestry, multi-traits and multi-methods. The three categories are not cumulative exhaustive or mutually exclusive. Multi-ancestry scores used multi-ancestry source of variant association or score development population. Multi-traits scores were identified by "incorporation of genetically correlated traits/CAD-related traits/risk factors" or similar wording in their source publications. Multi-methods scores refer to those developed by leveraging multiple scores of the same trait as made clear in source publications.

| PRS | Population Ranking | Individual Ranking | Method | Group | Score development sample size | Citation |
|---|---|---|---|---|---|---|
| **PGS000010** | 49 | 30 | Genome-wide significant variants | Ancestry-Specific | / | Mega JL *et al. Lancet* (2015) |
| **PGS000011** | 51 | 51 | Genome-wide significant variants | Multi-Ancestry | / | Tada H *et al. Eur Heart J* (2015) |
| **PGS000012** | 47 | 42 | LD thinning | Multi-Ancestry | 5883 | Abraham G *et al. Eur Heart J* (2016) |

| | | | | | | |
|---|---|---|---|---|---|---|
| **PGS000013** | 21 | 22 | LDpred | Multi-Ancestry | 120280 | Khera AV *et al. Nat Genet* (2018) |
| **PGS000018** | 19 | 28 | metaGRS | Multi-Ancestry, Multi-Method | 3000 | Inouye M *et al. J Am Coll Cardiol* (2018) |
| **PGS000019** | 43 | 44 | Genome-wide significant variants | Multi-Ancestry | / | Paquette M *et al. J Clin Lipidol* (2017) |
| **PGS000057** | 52 | 57 | Hard thresholding | Multi-Ancestry | / | Natarajan P *et al. Circulation* (2017) |
| **PGS000058** | 17 | 10 | Genome-wide significant variants | Multi-Ancestry | / | Morieri ML *et al. Diabetes Care* (2018) |
| **PGS000059** | 44 | 48 | Literature-derived SNP selection | Multi-Ancestry | / | Hajek C *et al. Circ Genom Precis Med* (2018) |
| **PGS000200** | 56 | 56 | Genome-wide significant variants | Multi-Ancestry | / | Tikkanen E *et al. Arterioscler Thromb Vasc Biol* (2013) |
| **PGS000296** | 25 | 36 | LDpred | Multi-Ancestry | 8766 | Wang M *et al. J Am Coll Cardiol* (2020) |
| **PGS000329** | 31 | 19 | LDpred | Ancestry-Specific | 21813 | Mars N *et al. Nat Med* (2020) |

| | | | | | | |
|---|---|---|---|---|---|---|
| **PGS000337** | 29 | 11 | P-value thresholding | Multi-Ancestry | / | Koyama S *et al. Nat Genet* (2020) |
| **PGS000349** | 57 | 58 | Genome-wide significant variants | Multi-Ancestry | / | Pechlivanis S *et al. BMC Med Genet* (2020) |
| **PGS000746** | 20 | 12 | LD clumping | Multi-Ancestry | 10000 | Gola D *et al. Circ Genom Precis Med* (2020) |
| **PGS000747** | 46 | 41 | LD clumping | Multi-Ancestry | 10000 | Gola D *et al. Circ Genom Precis Med* (2020) |
| **PGS000748** | 42 | 37 | Genome-wide significant variants | Multi-Ancestry | 10000 | Gola D *et al. Circ Genom Precis Med* (2020) |
| **PGS000749** | 39 | 31 | Genome-wide significant variants | Multi-Ancestry | 10000 | Gola D *et al. Circ Genom Precis Med* (2020) |
| **PGS000798** | 35 | 40 | Genome-wide significant variants | Ancestry-Specific | / | Severance LM *et al. J Cardiovasc Comput Tomogr* (2019) |
| **PGS000818** | 40 | 45 | Genome-wide significant variants | Multi-Ancestry | / | Bauer A *et al. Genet Epidemiol* (2021) |

| | | | | | | |
|---|---|---|---|---|---|---|
| **PGS000899** | 28 | 20 | SNP selection based on 1) genome-wide significance; 2) linkage disequilibrium | Multi-Ancestry | / | Feitosa MF *et al. Circ Genom Precis Med* (2021) |
| **PGS000962** | 37 | 32 | snpnet | Ancestry-Specific | 269704 | Tanigawa Y *et al. PLoS Genet* (2022) |
| **PGS001355** | 18 | 17 | AnnoPred | Ancestry-Specific | 92928 | Ye Y *et al. Circ Genom Precis Med* (2021) |
| **PGS001780** | 22 | 27 | PRS-CS | Multi-Ancestry | / | Tamlander M *et al. Commun Biol* (2022) |
| **PGS001839** | 11 | 13 | Penalize regression (bigstatsr) | Ancestry-Specific | 391124 | Privé F *et al. Am J Hum Genet* (2022) |
| **PGS002048** | 13 | 7 | LDpred2 (bigsnpr) | Ancestry-Specific | 391124 | Privé F *et al. Am J Hum Genet* (2022) |
| **PGS002244** | 30 | 18 | LDpred | Multi-Ancestry | 175364 | Mars N *et al. Cell Genom* (2022) |
| **PGS002262** | 54 | 49 | metaPRS | Multi-Ancestry, Multi-Trait | 4855 | Lu X *et al. Eur Heart J* (2022) |
| **PGS002775** | 24 | 29 | Maximum clumping and thresholding (maxCT) | Multi-Ancestry | 38217 | Wong CK *et al. PLoS One* (2022) |

| | | | | | | |
|---|---|---|---|---|---|---|
| **PGS002776** | 15 | 23 | Stacked clumping and thresholding (SCT) | Multi-Ancestry | 38217 | Wong CK *et al. PLoS One* (2022) |
| **PGS002809** | 38 | 35 | Genome-wide significant SNPs | Multi-Ancestry | / | Ahmed R *et al. Int J Cardiol Heart Vasc* (2022) |
| **PGS003355** | 16 | 25 | LDpred | Multi-Ancestry | / | Aragam KG *et al. Nat Genet* (2022) |
| **PGS003356** | 2 | 3 | LDpred | Multi-Ancestry | / | Aragam KG *et al. Nat Genet* (2022) |
| **PGS003438** | 6 | 9 | Genome-wide significant SNPs | Multi-Ancestry | / | Marston NA *et al. JAMA Cardiol* (2023) |
| **PGS003446** | 34 | 43 | PRSice | Multi-Ancestry | / | Tcheandjieu C *et al. Nat Med* (2022) |
| **PGS003725** | 4 | 4 | LDpred2 | Multi-Ancestry | 116649 | Patel AP *et al. Nat Med* (2023) |
| **PGS003726** | 3 | 1 | LDpred2 | Multi-Ancestry | 116649 | Patel AP *et al. Nat Med* (2023) |
| **PGS003727** | 1 | 2 | LDpred2 | Ancestry-Specific | 116649 | Patel AP *et al. Nat Med* (2023) |
| **PGS003866** | 12 | 15 | lassosum2 | Ancestry-Specific | / | Shim I *et al. Nature Communications* (2023) |

| | | | | | | |
|---|---|---|---|---|---|---|
| **PGS004196** | 26 | 39 | LASSO | Ancestry-Specific | 200000 | Raben TG *et al. Sci Rep* (2023) |
| **PGS004197** | 33 | 33 | LASSO | Ancestry-Specific | 200000 | Raben TG *et al. Sci Rep* (2023) |
| **PGS004198** | 36 | 47 | LASSO | Ancestry-Specific | 200000 | Raben TG *et al. Sci Rep* (2023) |
| **PGS004199** | 27 | 46 | LASSO | Ancestry-Specific | 200000 | Raben TG *et al. Sci Rep* (2023) |
| **PGS004200** | 32 | 38 | LASSO | Ancestry-Specific | 200000 | Raben TG *et al. Sci Rep* (2023) |
| **PGS004321** | 45 | 24 | Genome-wide significant SNPs | Multi-Ancestry | / | Marston NA *et al. Circulation* (2019) |
| **PGS004443** | 41 | 26 | LDpred2 | Ancestry-Specific | 174489 | Jung H *et al. Commun Biol* (2024) |
| **PGS004444** | 55 | 55 | LDpred2 | Ancestry-Specific | 174489 | Jung H *et al. Commun Biol* (2024) |
| **PGS004513** | 53 | 50 | RFDiseasemetaPRS | Ancestry-Specific, | 174489 | Jung H *et al. Commun Biol* (2024) |

| | | | | | | |
|---|---|---|---|---|---|---|
| | | | | Multi-Trait | | |
| **PGS004514** | 58 | 52 | RFDiseasemetaPRS | Ancestry-Specific, Multi-Trait | 174489 | Jung H *et al. Commun Biol* (2024) |
| **PGS004595** | 48 | 53 | Genome-wide significant SNPs | Ancestry-Specific | / | Oni-Orisan A *et al. Clin Pharmacol Ther* (2022) |
| **PGS004596** | 50 | 54 | Genome-wide significant SNPs | Multi-Ancestry | / | Peng H *et al. Nutrients* (2023) |
| **PGS004696** | 7 | 6 | PRS-CSx | Multi-Ancestry | 87724 | Smith JL *et al. Circ Genom Precis Med* (2024) |
| **PGS004697** | 5 | 5 | PRS-CSx | Multi-Ancestry | 56359 | Smith JL *et al. Circ Genom Precis Med* (2024) |
| **PGS004698** | 8 | 16 | Pruning and Thresholding | Multi-Ancestry | 87724 | Smith JL *et al. Circ Genom Precis Med* (2024) |
| **PGS004743** | 9 | 8 | PRSmix | Ancestry-Specific, Multi-Method | 29863 | Truong B et al., *Cell Genom* (2024) |
| **PGS004744** | 14 | 34 | PRSmix | Ancestry-Specific, | 35350 | Truong B et al., *Cell Genom* (2024) |

| | | | | | | |
|---|---|---|---|---|---|---|
| | | | | Multi-Method | | |
| **PGS004745** | 23 | 21 | PRSmix | Ancestry-Specific, Multi-Method | 29863 | Truong B et al., *Cell Genom* (2024) |
| **PGS004746** | 10 | 14 | PRSmixPlus | Ancestry-Specific, Multi-Method | 35350 | Truong B et al., *Cell Genom* (2024) |

Information on scores (method, score development sample size and citation) extracted from PGS catalog (https://www.pgscatalog.org).

**Supplementary Table 11. Summary of evaluated CAD PRS scores not using UKBB genome-wide association study (GWAS) data in the primary analysis.** GWAS data of PRS scores extracted from PGS catalog (https://www.pgscatalog.org).

| PRS | Population Ranking | Individual Ranking | Method | Group | Score development sample size | Citation |
|---|---|---|---|---|---|---|
| **PGS000010** | 26 | 26 | Genome-wide significant variants | Ancestry-Specific | / | Mega JL *et al. Lancet* (2015) |
| **PGS000011** | 27 | 28 | Genome-wide significant variants | Multi-Ancestry | / | Tada H *et al. Eur Heart J* (2015) |
| **PGS000012** | 22 | 24 | LD thinning | Multi-Ancestry | 5883 | Abraham G *et al. Eur Heart J* (2016) |
| **PGS000013** | 12 | 11 | LDpred | Multi-Ancestry | 120280 | Khera AV *et al. Nat Genet* (2018) |
| **PGS000018** | 10 | 10 | metaGRS | Multi-Ancestry, Multi-Method | 3000 | Inouye M *et al. J Am Coll Cardiol* (2018) |
| **PGS000019** | 21 | 22 | Genome-wide significant variants | Multi-Ancestry | / | Paquette M *et al. J Clin Lipidol* (2017) |

| | | | | | | |
|---|---|---|---|---|---|---|
| **PGS000057** | 28 | 27 | Hard thresholding | Multi-Ancestry | / | Natarajan P *et al.* *Circulation* (2017) |
| **PGS000059** | 25 | 21 | Literature-derived SNP selection | Multi-Ancestry | / | Hajek C *et al. Circ Genom Precis Med* (2018) |
| **PGS000200** | 29 | 29 | Genome-wide significant variants | Multi-Ancestry | / | Tikkanen E *et al. Arterioscler Thromb Vasc Biol* (2013) |
| **PGS000296** | 14 | 15 | LDpred | Multi-Ancestry | 8766 | Wang M *et al. J Am Coll Cardiol* (2020) |
| **PGS000349** | 30 | 30 | Genome-wide significant variants | Multi-Ancestry | / | Pechlivanis S *et al. BMC Med Genet* (2020) |
| **PGS000746** | 16 | 17 | LD clumping | Multi-Ancestry | 10000 | Gola D *et al. Circ Genom Precis Med* (2020) |
| **PGS000747** | 17 | 14 | LD clumping | Multi-Ancestry | 10000 | Gola D *et al. Circ Genom Precis Med* (2020) |
| **PGS000748** | 20 | 20 | Genome-wide significant variants | Multi-Ancestry | 10000 | Gola D *et al. Circ Genom Precis Med* (2020) |
| **PGS000749** | 19 | 19 | Genome-wide significant variants | Multi-Ancestry | 10000 | Gola D *et al. Circ Genom Precis Med* (2020) |

| | | | | | | |
|---|---|---|---|---|---|---|
| **PGS001780** | 11 | 12 | PRS-CS | Multi-Ancestry | / | Tamlander M *et al. Commun Biol* (2022) |
| **PGS002244** | 15 | 16 | LDpred | Multi-Ancestry | 175364 | Mars N *et al. Cell Genom* (2022) |
| **PGS002262** | 24 | 25 | metaPRS | Multi-Ancestry, Multi-Trait | 4855 | Lu X *et al. Eur Heart J* (2022) |
| **PGS002775** | 18 | 18 | Maximum clumping and thresholding (maxCT) | Multi-Ancestry | 38217 | Wong CK *et al. PLoS One* (2022) |
| **PGS002776** | 13 | 13 | Stacked clumping and thresholding (SCT) | Multi-Ancestry | 38217 | Wong CK *et al. PLoS One* (2022) |
| **PGS003355** | 9 | 6 | LDpred | Multi-Ancestry | / | Aragam KG *et al. Nat Genet* (2022) |
| **PGS003725** | 6 | 7 | LDpred2 | Multi-Ancestry | 116649 | Patel AP *et al. Nat Med* (2023) |
| **PGS003726** | 7 | 9 | LDpred2 | Multi-Ancestry | 116649 | Patel AP *et al. Nat Med* (2023) |
| **PGS003727** | 8 | 8 | LDpred2 | Ancestry-Specific | 116649 | Patel AP *et al. Nat Med* (2023) |
| **PGS003866** | 4 | 3 | lassosum2 | Ancestry-Specific | / | Shim I *et al. Nature Communications* (2023) |

| | | | | | | |
|---|---|---|---|---|---|---|
| **PGS004321** | 23 | 23 | Genome-wide significant SNPs | Multi-Ancestry | / | Marston NA *et al.* *Circulation* (2019) |
| **PGS004743** | 2 | 2 | PRSmix | Ancestry-Specific, Multi-Method | 29863 | Truong B et al., *Cell Genom* (2024) |
| **PGS004744** | 5 | 5 | PRSmix | Ancestry-Specific, Multi-Method | 35350 | Truong B et al., *Cell Genom* (2024) |
| **PGS004745** | 1 | 4 | PRSmix | Ancestry-Specific, Multi-Method | 29863 | Truong B et al., *Cell Genom* (2024) |
| **PGS004746** | 3 | 1 | PRSmixPlus | Ancestry-Specific, Multi-Method | 35350 | Truong B et al., *Cell Genom* (2024) |

Information on scores (method, score development sample size and citation) extracted from PGS catalog (https://www.pgscatalog.org).
