## Supplementary Figure 1-7 for "Evaluating Individual Level Performance of Polygenic Risk Scores Using Early Onset High Genetic Risk Coronary Artery Disease as a Benchmark"

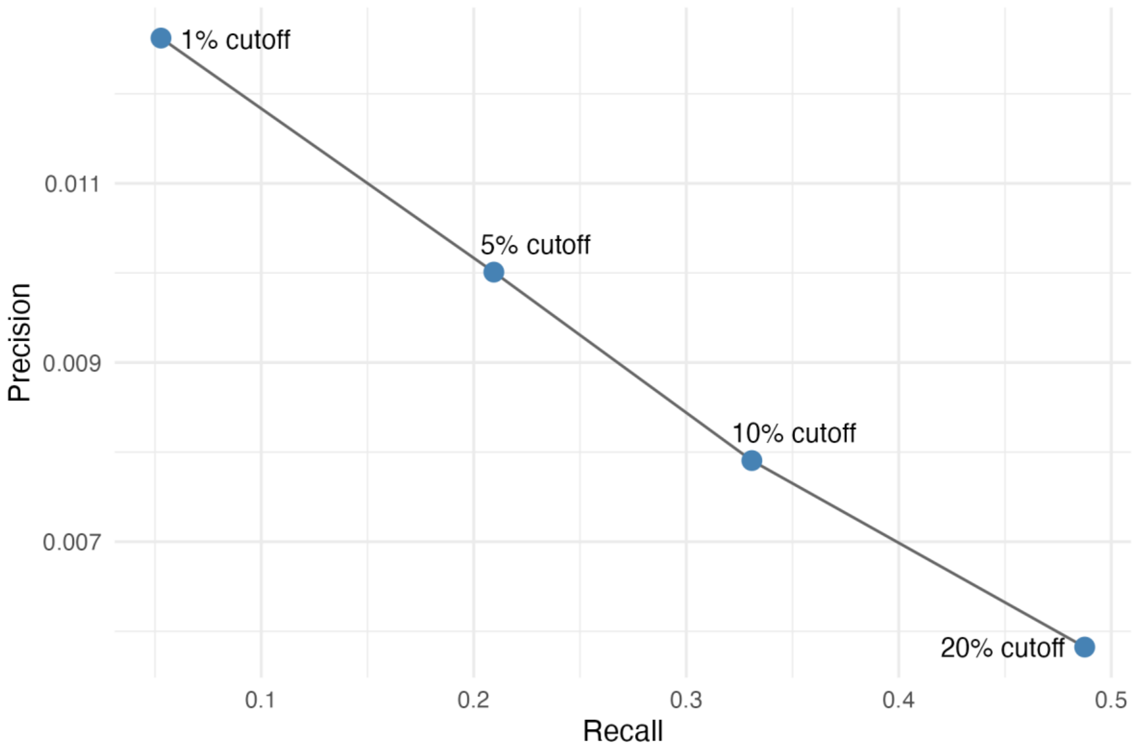


**Supplementary Figure 1. Precision-recall curve comparing various threshold of top PRS using PGS004697 as an example.** Recall is equivalent to sensitivity, or percentage of benchmark early-onset unexpected CAD cohort identified using various cutoffs as thresholds of high PRS. Precision is the positive predictive value, quantifying the proportion of patients from the benchmark cohort identified as high PRS with various cutoffs.

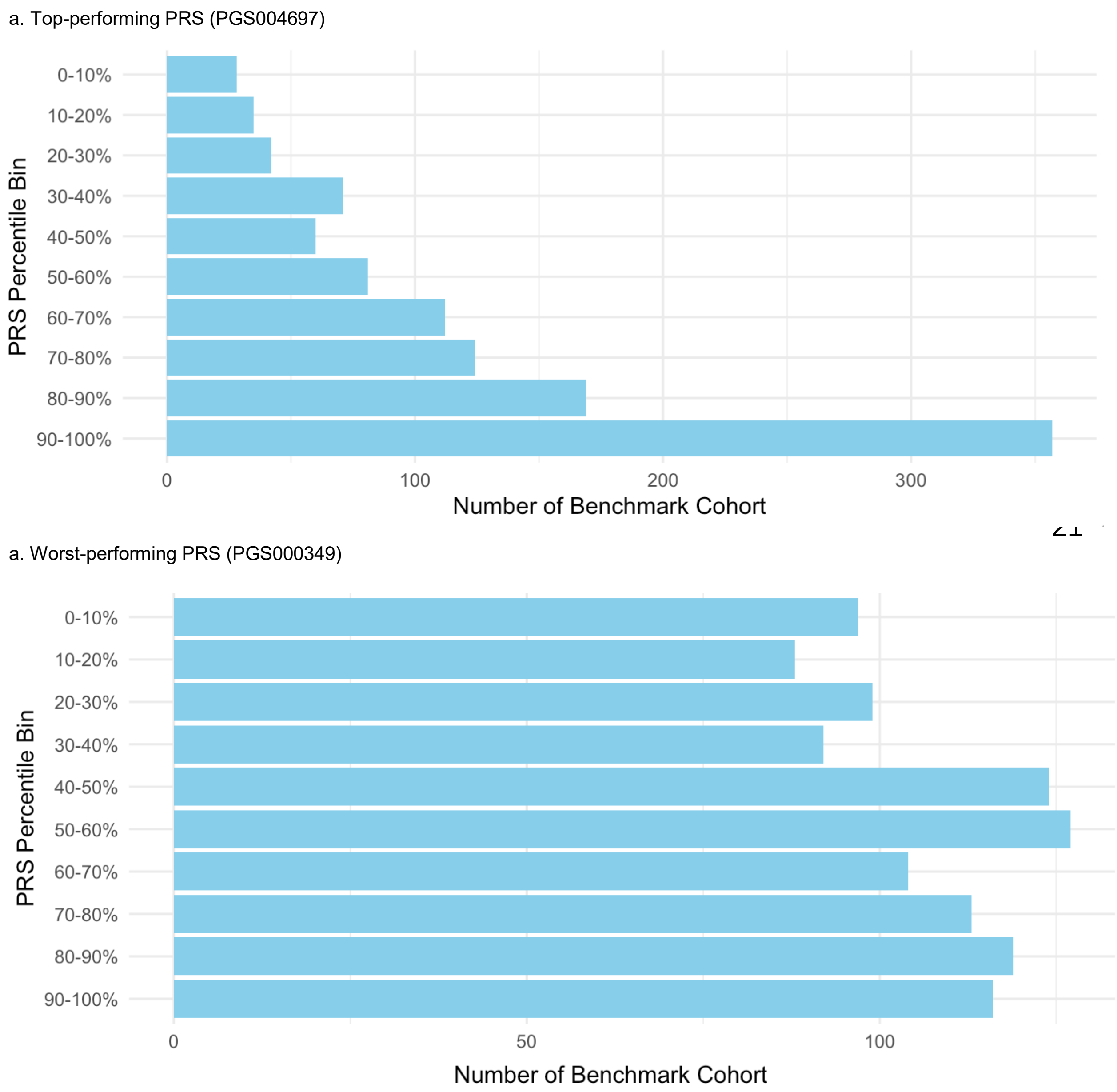


**Supplementary Figure 2. Distribution of early-onset unexpected CAD benchmark cohort across PRS deciles.** (a) When evaluating the top individual-level performing PRS (PGS004697), early-onset unexpected CAD benchmark cohort individuals were enriched in the top decile. (b) When evaluating the worst individual-level performing PRS (PGS000349), early-onset unexpected CAD benchmark cohort individuals were more uniformly distributed across PRS deciles.

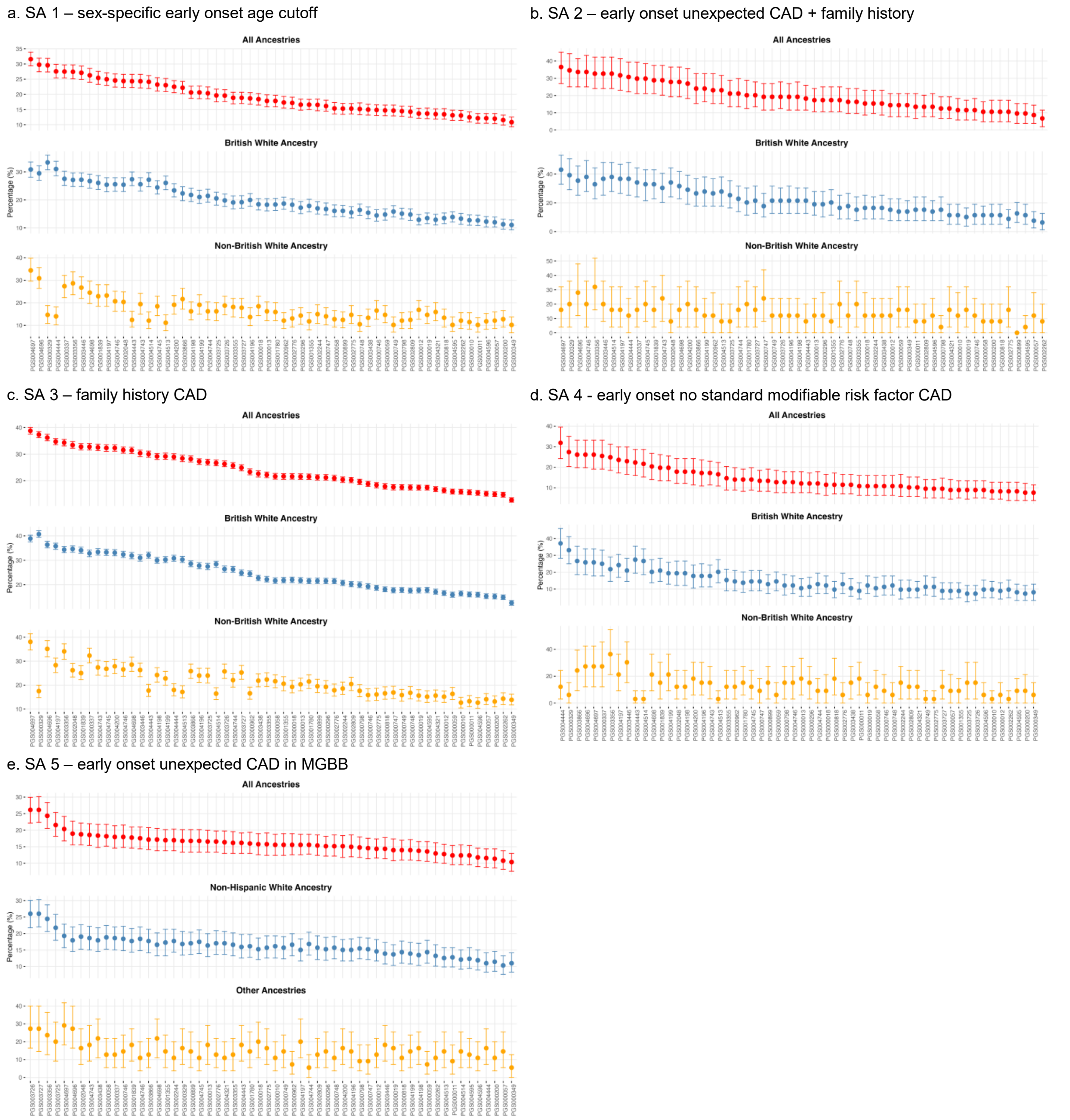


**Supplementary Figure 3. Individual-level risk prediction performance of 58 CAD PRS scores by ancestry in sensitivity analysis 1-5.** Individual-level risk prediction of PRS scores was measured by percentage of benchmark population within top decile of each PRS out of UKBB (sensitivity analysis 1-4, n=451,580) or MGBB (sensitivity analysis 5, n=49,744). The scores were ranked by decreasing percentage capture in all ancestries. Error bars represent 95% confidence intervals obtained via bootstrapping. (a) In sensitivity analysis

1, benchmark cohort was defined in the same way as the primary analysis, except for differential early onset cutoff age (60 years old) for females. (b) In sensitivity analysis 2, benchmark cohort was defined as those in the primary analysis benchmark cohort also having family history of heart disease in both parents. (c) In sensitivity analysis 3, benchmark cohort was defined as CAD patients with family history of heart disease in both parents. (d) In sensitivity analysis 4, benchmark cohort was defined as early onset CAD patients with no standard modifiable risk factors in UKBB. (e) In sensitivity analysis 5, benchmark cohort was defined as early onset CAD patients with no standard modifiable risk factors in MGBB.

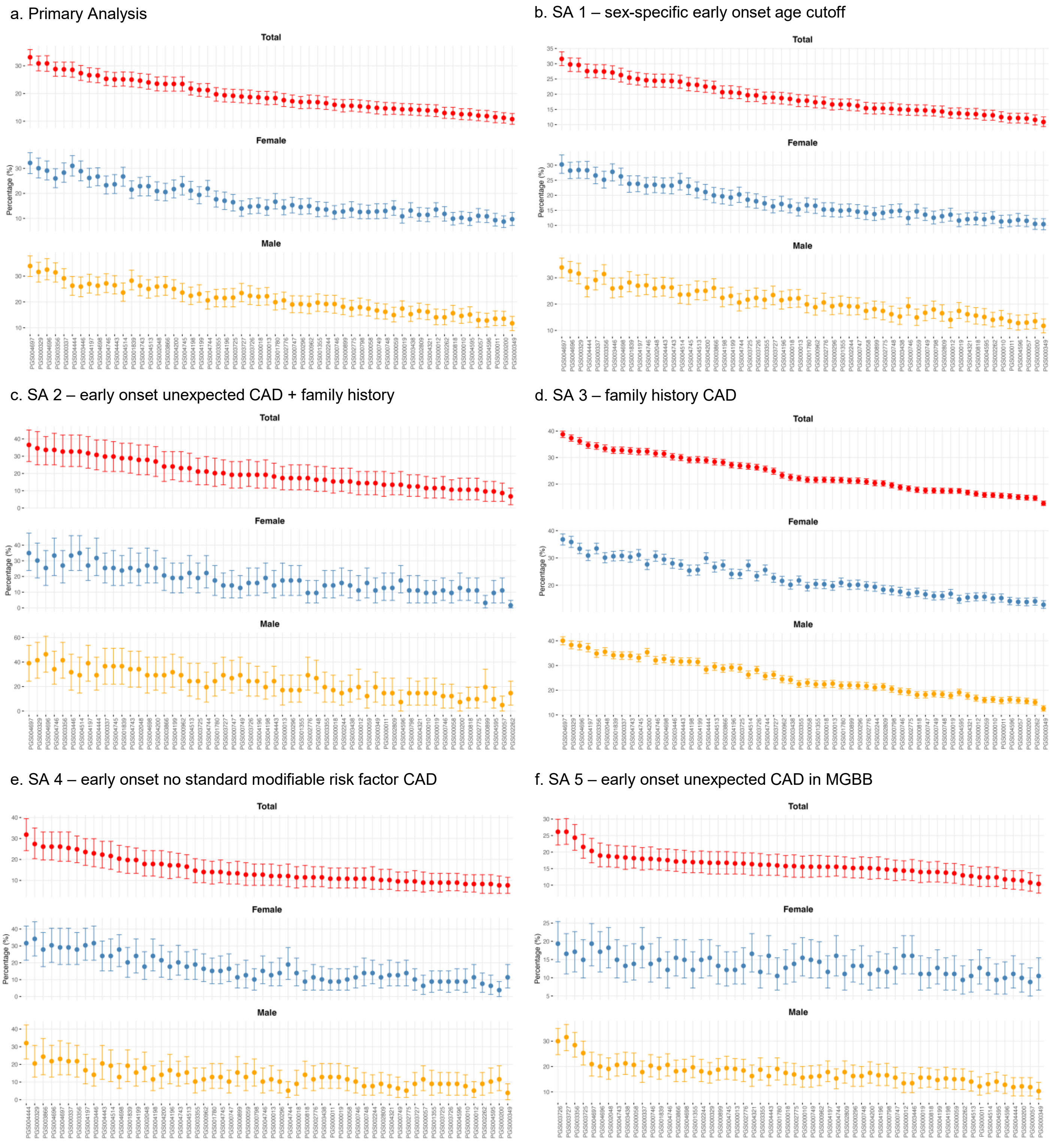


**Supplementary Figure 4. Individual-level risk prediction performance of 58 CAD PRS scores by sex in primary analysis and sensitivity analysis 1-5.** Individual-level risk prediction of PRS scores was measured by percentage of benchmark population within top decile of each PRS out of UKBB (primary analysis and sensitivity analysis 1-4, n=451,580) or MGBB (sensitivity analysis 5, n=49,744). The scores were ranked by decreasing percentage capture in total population. Error bars represent 95% confidence intervals obtained via bootstrapping. (a) In primary analysis, benchmark cohort was defined as early onset CAD patients with no standard indications for lipid lowering therapy in UKBB. (b) In sensitivity analysis 1, benchmark cohort was defined in the same way as the primary analysis, except for differential early onset cutoff age (60 years old) for females. (c) In sensitivity analysis 2, benchmark cohort was defined as those in the primary analysis benchmark cohort also having family history of heart disease in both parents. (d) In sensitivity analysis 3, benchmark cohort was defined as CAD patients with family history of heart disease in both parents. (e) In sensitivity analysis 4, benchmark cohort was defined as early onset CAD patients with no standard modifiable risk factors in UKBB. (f) In sensitivity analysis 5, benchmark cohort was defined as early onset CAD patients with no standard modifiable risk factors in MGBB.

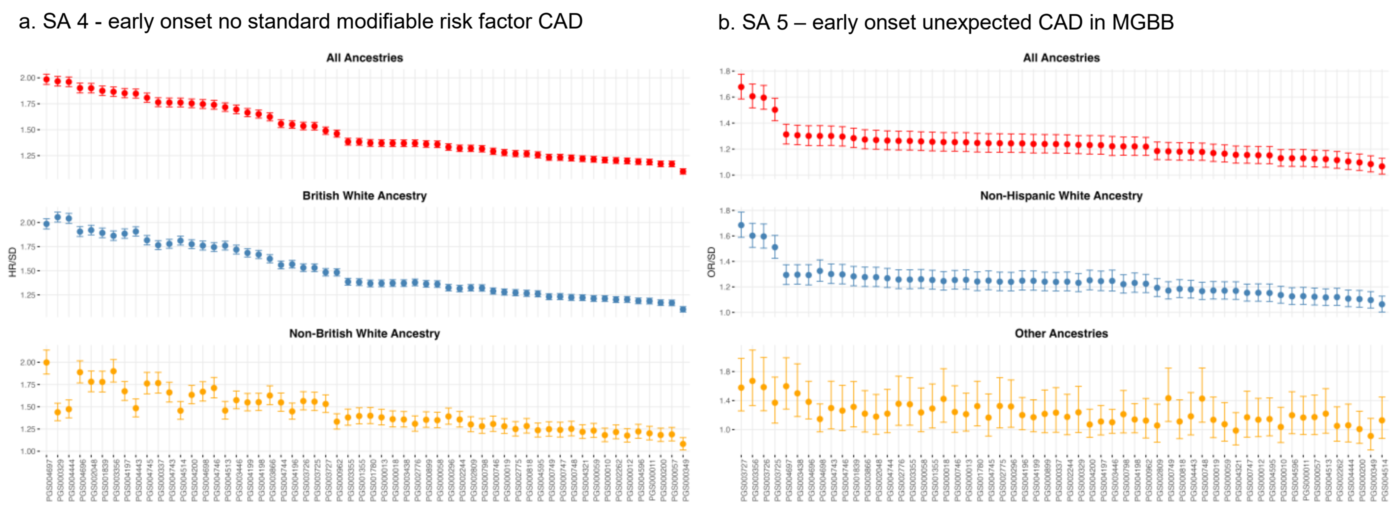


**Supplementary Figure 5. Population-level performance of 58 CAD PRS by ancestry in sensitivity analysis 4 and 5.** Error bars represent 95% confidence intervals obtained via bootstrapping. (a) In sensitivity analysis 4, population-level performance of PRS scores was conventionally assessed with hazard ratio (HR) per one standard deviation increase in PRS using a Cox proportional hazard model in UKBB population with no prevalent CAD or risk factors (hypertension, diabetes mellitus, hypercholesterolemia). The model censored individuals at the time of developing any of the risk factors and was adjusted for age, sex and PC1-PC4. The scores were ranked by decreasing HR in all ancestries. (b) In sensitivity analysis 5, population-level performance of PRS scores was conventionally assessed with odds ratio (OR) per one standard deviation increase in PRS using a logistic regression model in MGBB population. The model was adjusted for age, sex and categorical ancestry. The scores were ranked by decreasing OR in all ancestries.

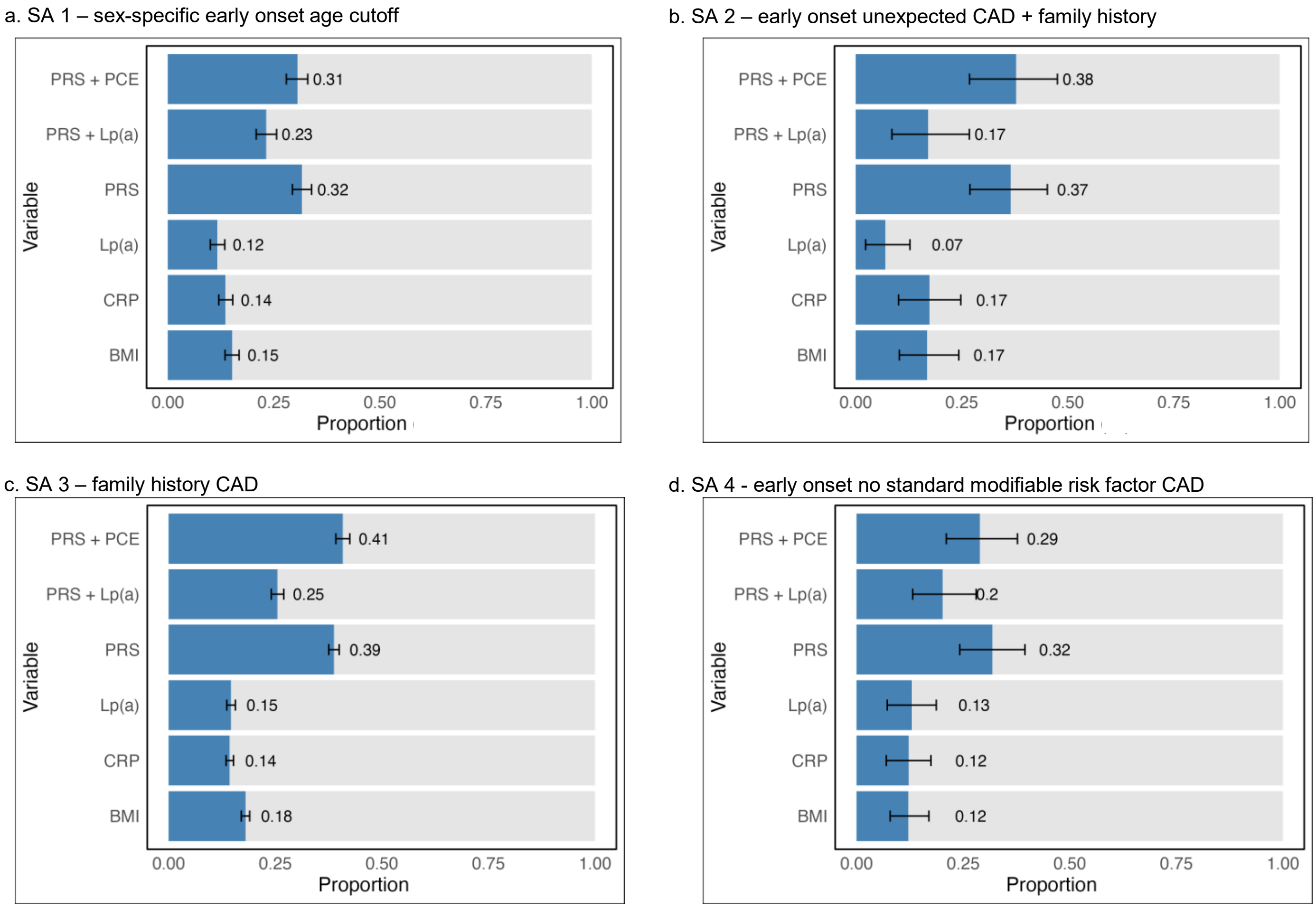


**Supplementary Figure 6. Proportion of benchmark CAD patients captured at the top decile of PRS versus other clinical biomarkers.** The PRS used for comparison in each analysis is the best-performing PRS among the 58 CAD PRS evaluated in the corresponding analysis. Lp(a) was standardized before combining with PRS. Error bars represent 95% confidence intervals obtained via bootstrapping. (a) In sensitivity analysis 1, benchmark cohort was defined in the same way as the primary analysis, except for differential early onset cutoff age (60 years old) for females. (b) In sensitivity analysis 2, benchmark cohort was defined as those in the primary analysis benchmark cohort also having family history of heart disease in both parents. (c) In sensitivity analysis 3, benchmark cohort was defined as CAD patients with family history of heart disease in both parents. (d) In sensitivity analysis 4, benchmark cohort was defined as early onset CAD patients with no standard modifiable risk factors in UKBB. BMI = body mass index; CRP = C-reactive protein; Lp(a) = lipoprotein (a); PCE = Pooled Cohort Equation; PRS = polygenic risk score.

a. SA 1 – sex-specific early onset age cutoff

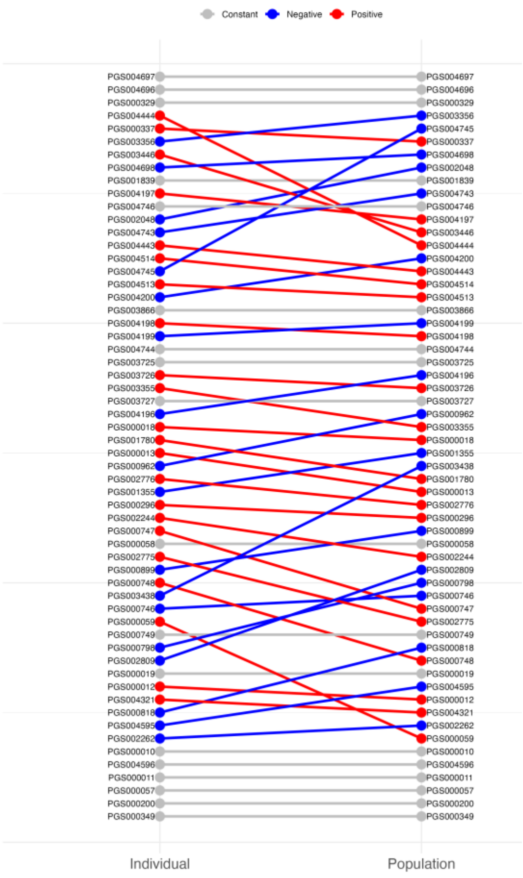


b. SA 2 – early onset unexpected CAD + family history

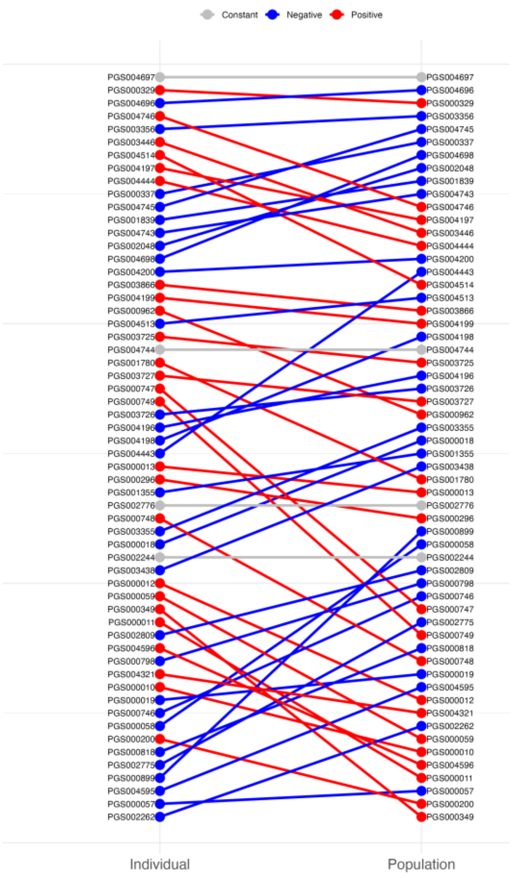


c. SA 3 – family history CAD

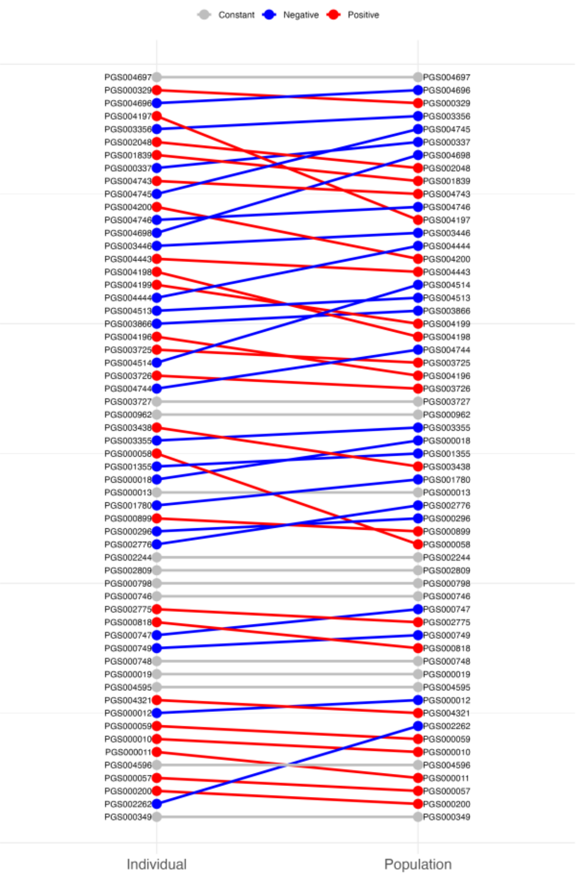


d. SA 4 - early onset no standard modifiable risk factor CAD

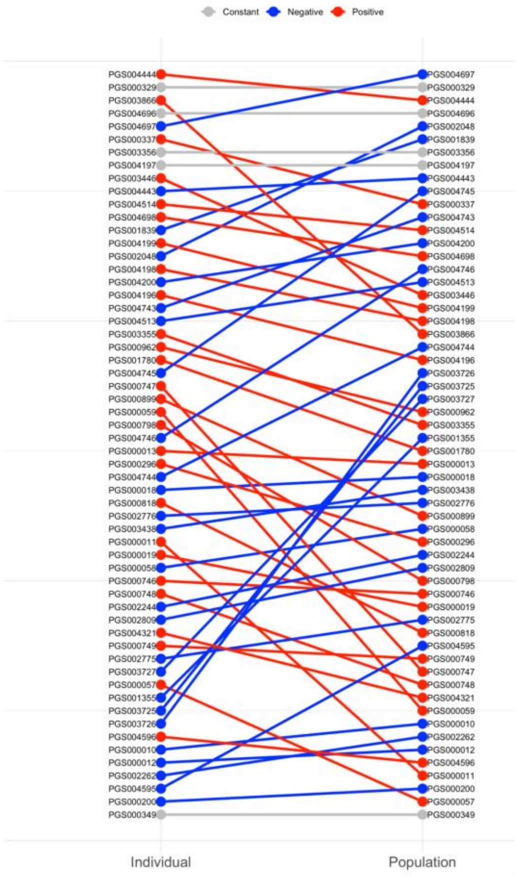


e. SA 5 – early onset unexpected CAD in MGBB

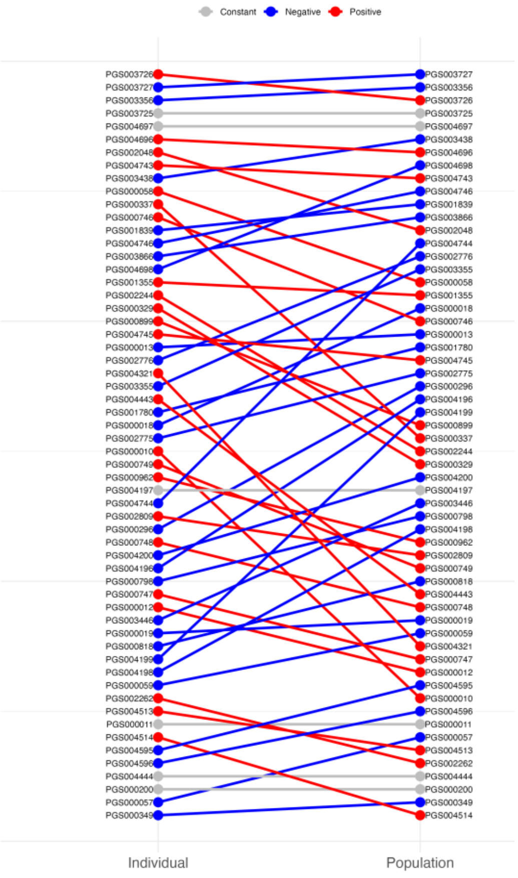


f. Ranking changes using 5% as high PRS threshold

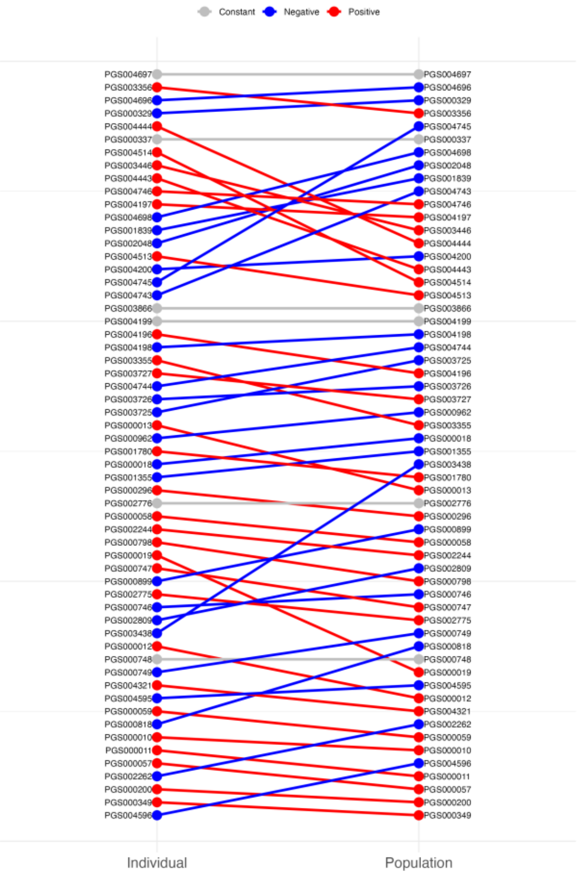

**Supplementary Figure 7. Rank transition of CAD PRSs at individual-level and population-level performance spaces in sensitivity analysis 1-5 (a-e) and using 5% as high PRS threshold (f).** Red indicates positive slope: higher individual-level than population-level performance ranking. Blue indicates negative slope: higher population-level than individual-level performance ranking. Grey indicates constant slope and same performance ranking. (a) In sensitivity analysis 1, benchmark cohort was defined in the same way as the primary analysis, except for differential early onset cutoff age (60 years old) for females. (b) In sensitivity analysis 2, benchmark cohort was defined as those in the primary analysis benchmark cohort also having family history of heart disease in both parents. (c) In sensitivity analysis 3, benchmark cohort was defined as CAD patients with family history of heart disease in both parents. (d) In sensitivity analysis 4, benchmark cohort was defined as early onset CAD patients with no standard modifiable risk factors in UKBB. (e) In sensitivity analysis 5, benchmark cohort was defined as early onset CAD patients with no standard modifiable risk factors in MGBB. (f) Using 5% instead of 10% in primary analysis as high PRS threshold, PRS ranking by individual-level metric highly correlated with that of the primary analysis (Spearman’s rho = 0.98).
